# Longitudinal Hierarchical Bayesian models of covariate effects on airway and alveolar nitric oxide

**DOI:** 10.1101/2022.10.12.22281006

**Authors:** Jingying Weng, Noa Molshatzki, Paul Marjoram, W. James Gauderman, Frank D. Gilliland, Sandrah P. Eckel

## Abstract

Biomarkers such as exhaled nitric oxide (FeNO), a marker of airway inflammation, have applications in the study of chronic respiratory disease where longitudinal studies of within-participant changes in the biomarker are particularly relevant. A cutting-edge approach to assessing FeNO, called multiple flow FeNO, repeatedly assesses FeNO across a range of expiratory flow rates at a single visit and combines these data with a deterministic model of lower respiratory tract NO to estimate parameters quantifying airway wall and alveolar NO sources. Previous methodological work for multiple flow FeNO has focused on methods for data from a single participant or from cross-sectional studies. Performance of existing *ad hoc* two-stage methods for longitudinal multiple flow FeNO in cohort or panel studies has not been evaluated. In this paper, we present a novel longitudinal extension to a unified hierarchical Bayesian (L_U_HB) model relating longitudinally assessed multiple flow FeNO to covariates. In several simulation study scenarios, we compare the L_U_HB method to other unified and two-stage frequentist methods. In general, L_U_HB produced unbiased estimates, had good power, and its performance was not sensitive to the magnitude of the association with a covariate and correlations between NO parameters. In an application relating height to longitudinal multiple flow FeNO in schoolchildren without asthma, unified analysis methods estimated positive, statistically significant associations of height with airway and alveolar NO concentrations and negative associations with airway wall diffusivity while estimates from two-stage methods were smaller in magnitude and sometimes non-significant.

## Introduction

Sophisticated statistical methods are needed to link longitudinal assessments of a biomarker to patient-level characteristics and time-varying exposures in the context of a deterministic mathematical model describing the production and dynamics of the biomarker within the human body. This paper presents statistical methods developed for such longitudinal assessments of the fractional concentration of exhaled nitric oxide, FeNO, a biomarker of airway inflammation used in clinical [1-3] and epidemiological research[4-6].

FeNO is an exhaled breath biomarker conventionally assessed at the target expiratory flow rate of 50 ml/s (FeNO_50_)[7]. A cutting-edge approach, called multiple flow FeNO, repeatedly assesses FeNO across a range of expiratory flow rates and combines these data with a deterministic model of NO in the lower respiratory tract to estimate parameters quantifying the effect of airway wall and alveolar sources. Literature on the modeling of multiple flow FeNO data has focused on methods for data collected from one person at a single visit[8-11]. Multiple flow FeNO data present statistical challenges which require sophisticated statistical methods. Many studies of multiple flow FeNO conduct analyses using a two-stage approach: (1) estimate airway and alveolar NO parameters and (2) treat the estimated NO parameters as observed outcomes in linear regressions relating NO parameters to factors of interest (asthma medication use, air pollution exposures, etc.). In a previous paper, we presented a novel unified hierarchical Bayesian (U-HB) model for estimating *cross-sectional* associations of covariates with NO parameters using data from a single multiple flow FeNO test session for each study participant. We also found, in an extensive simulation study, that the U_HB method was less biased and had better power/type I error compared to conventional two-stage methods[12].

There is a need for longitudinal data analysis methods for FeNO. Biomarkers like FeNO are particularly promising for tracking within-person changes over time since they tend to be relatively stable within persons, despite considerable heterogeneity across people[13-16]. Longitudinal trends in study populations with repeated measures of the conventional FeNO_50_ are generally modeled using standard longitudinal data analysis techniques, such as linear mixed effects models (LMM) or generalized additive linear mixed effects models (GAMM) [17]. Panel studies or longitudinal cohort studies with repeated measures of multiple flow FeNO across multiple visits can highlight within-person trends in proximal and distal inflammation. However, there have been no methods proposed for longitudinal multiple flow FeNO data, and the performance of existing *ad hoc* two-stage methods has not been evaluated.

Here, we present a novel extension of the U-HB model to longitudinal data (L-U-HB) using a Bayesian implementation of nonlinear mixed effects models. L-U-HB takes as input longitudinal measurements of multiple-flow FeNO on a group of participants to estimate associations of NO parameters with time-varying or time-constant covariates, as well as to quantify within- and between-participant variation. Our work is motivated by longitudinal multiple flow FeNO data collected as part of the Southern California Children’s Health Study (CHS) [18]. The CHS, originally designed to study impacts of long-term air pollution exposures on children’s respiratory health, included repeated measurements of multiple flow FeNO in the most recent cohort.

## Methods

First, we introduce the mathematical deterministic model for FeNO in the respiratory tract and then we describe statistical methods for estimating NO parameters from this model using multiple flow FeNO data, including two-stage (TS) approaches and unified (U) approaches for longitudinally assessed multiple flow FeNO.

### Deterministic two compartment model for FeNO

Our work is based on the simple steady-state two-compartment model (2CM), which assumes a cylindrically-shaped airway compartment with related NO parameters: *C*_*aw*_, the concentration of NO in the airway tissue (ppb); *D*_*aw*_, the airway tissue diffusion capacity (pL·s^−1^·ppb^−1^), and an expansile alveolar compartment with related NO parameter *C*_*A*_, the concentration of NO in the alveolar region (ppb) [8]. Under the 2CM, FeNO (ppb) at the mouth is deterministically related to expiratory flow rate (ml/s) and the three NO parameters quantifying airway and alveolar sources of NO, as shown below:

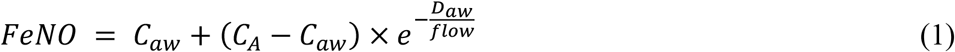

### Estimating NO parameters in the 2CM

The 2CM model for FeNO in Equation 1 is deterministic and nonlinear. In practice, multiple flow FeNO data is measured with error. Researchers have developed various methods to estimate 2CM NO parameters using multiple flow data from a given participant, typically using linear regression approaches with an underlying linearization assumption, a third order approximation method such as the Högman and MerilLinen algorithm (HMA) [19, 20], and nonlinear regression which essentially adds an error term to the right hand side of Equation 1 [21]. Here and in previous work[10, 12], we use the following fundamental nonlinear statistical model for multiple flow FeNO measured repeatedly across a range of flow rates for a single participant, with maneuvers indexed by *k*:

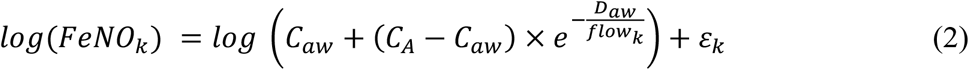

This model formulation includes a “transform-both-sides” [22] approach using the natural log to acknowledge the increased variation in error that occurs as flow rate (and hence FeNO concentration) increases w hile maintaining the interpretability of the 2CM NO parameters. On the logFeNO scale, the error (ε) can be reasonably assumed to be normally distributed. Henceforth, we will refer to a model estimating NO parameters using Equation 2 with standard nonlinear-least squares software (e.g., “nls” from the nlme package in R) as NLS [10]. So far, we have discussed only estimation of NO parameters from a single multiple flow FeNO test session for one participant. When multiple flow FeNO data are assessed longitudinally in a study population the data have three levels of variation: across-participant, within-participant (across visits), and within-visit (across maneuvers).

### Estimating associations of covariates with longitudinally assessed NO parameters Two Stage (TS) methods

In the existing literature, most researchers use *ad hoc* two stage approaches to relate estimated NO parameters to covariates. Two stage methods for cross-sectional studies were discussed in our previous work[12]. A typical longitudinal two stage method proceeds as follows. In Stage I, NO parameters for each participant at each visit are estimated via separate models. For example, a separate HMA or NLS model is fit to the multiple flow FeNO data from each participant at each visit. In Stage II, three linear mixed effect models (LMMs) are fit, one for each NO parameter, to relate the longitudinal estimates of the NO parameters to a covariate(s) of interest, denoted generically as X_ij_. A participant-level random intercept is included in each LMM to account for the within-participant correlation in the longitudinal NO parameter data. Below, we introduce 4 longitudinal two stage (L_TS) methods, differentiated by the name of the method employed in Stage I:

1. L_TS_NLS: Stage I consists of N (participants) x M (visits) separate NLS [10] models, each NLS model fit to the typically small multiple flow FeNO dataset at that visit.
2. L_TS_HMA: Similarly, Stage I consists of N x M separate HMA [19, 20] models.
3. L_TS_NLME: Stage I consists of a single longitudinal nonlinear least square mixed effect (NLME) model, an extension of the approach using N x M separate NLS models. In the longitudinal NLME, we specified participant-level and visit-level random intercepts for each NO parameter. At each level, these random effects follow a multivariate normal distribution, allowing for correlation of NO parameters. For example, participant-level correlation allows for participants with high C_A_ to also tend to have high *C*_*aw*_. We implemented NLME using the nlme package in R (version 3.1-152) [23].
4. L_TS_HB: Stage I consists of a single longitudinal Hierarchical Bayesian (HB) analog of the longitudinal NLME model, implemented using JAGS (Just another Gibbs sampler [24]) similar to the U-HB cross-sectional model in our previous publication[12], but with no covariate X and a partitioning of variance in NO parameters at the participant- and visit-levels through specification of variance-covariance matrices, the population mean of the NO parameters, and the measurement error in Stage I. This model is described in greater detail in the L-U-HB section below, where the model includes X.

All these TS approaches use the same LMM approach in Stage II.

### Unified approaches

Unified methods, in contrast to TS methods, simultaneously estimate NO parameters and their associations with the covariate *X*_*ij*_ in a single model. In this longitudinal version, we estimated the between/within-participant variation at the same time, which L-TS-NLS and L-TS-HMA were not able to obtain because their estimation in the stage I ignored the grouping effect.

#### 5. L-U-HB

Our novel U-HB model for longitudinal data (U-HBL) has three levels: maneuver, visit, participants, as described below and displayed in Figure 1:

**Figure 1:**
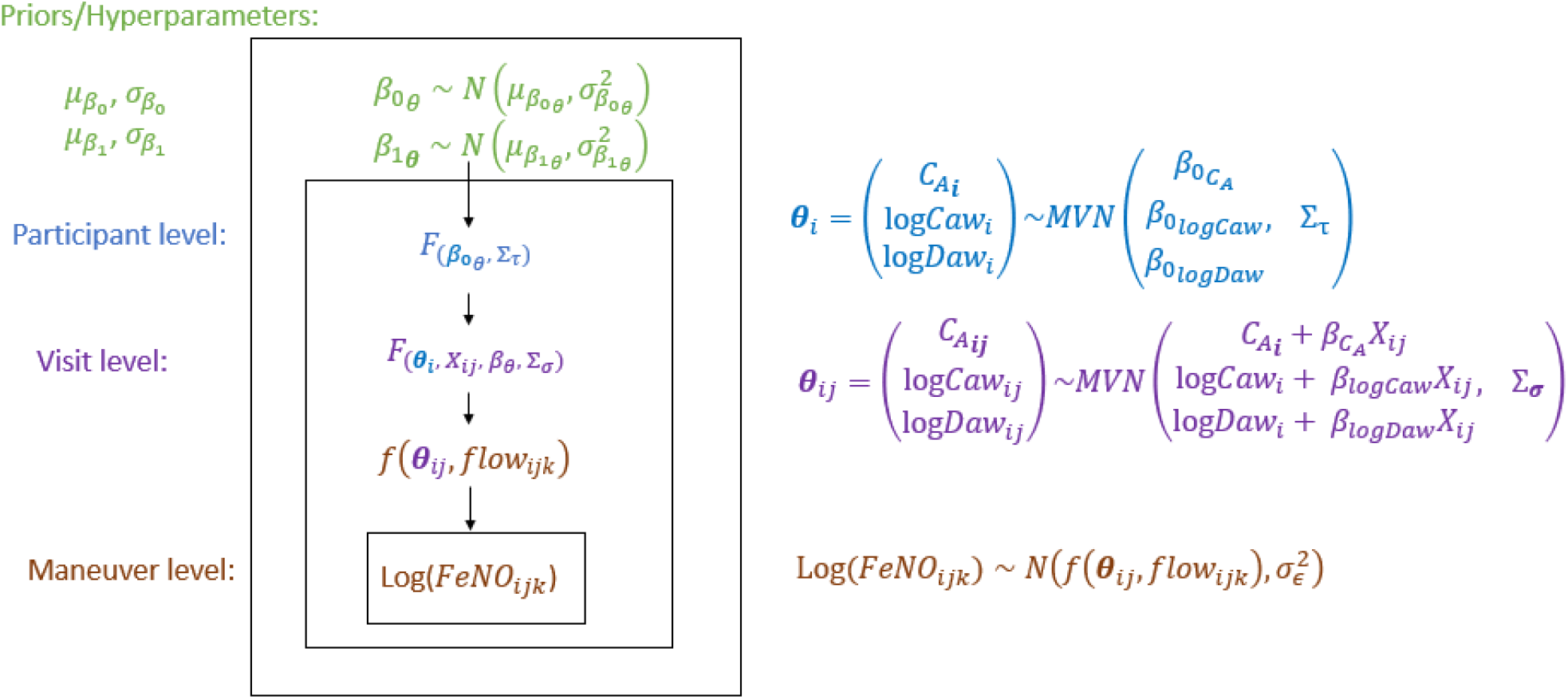
Diagram of the hierarchical model structure relating Longitudinal FeNO measurements at multiple flow rates to NO parameters that are a function of a potential determinant X (e.g., air pollution)

Level 1: Maneuver

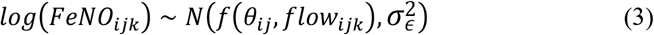

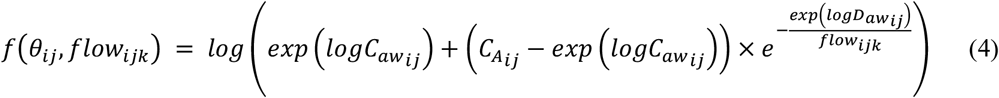

In the first level, log FeNO for participant *i* at visit *j* and maneuver *k* is assumed to be normally distributed with a mean that is a function of NO parameters: *θ*ij =(*C*_*Aij*_, *logC*_*awij*_, *logD*_*awij*_) ′ and expiratory flow, *flow*_*ijk*_. The variance of the unexplained error in logFeNO, 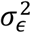, was assumed to be the same across flow rates, visits, and participants.

Level 2: Visit (time)

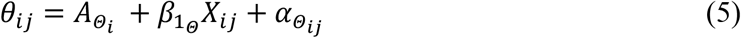

In the second level, NO parameters for participant *i* at visit *j* (*θ*_*ij*_) are modeled as a linear function of 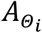, a vector of participant-level mean NO parameter values for participant *i* when the covariate *X*_*ij*_ =0. Key parameters of interest include *β* _*Θ*_= (*β*_*ca*_, *β*_*logCaw*_, *β*_*logDaw*_)′, the regression coefficients on *X*_*ij*_. Otherwise unexplained within-participant variation in the NO parameters is represented by the visit-level random intercepts 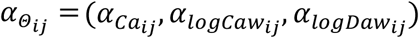 assumed to have no correlations and follow a multivariate normal distribution (MVN) with variance-covariance matrix 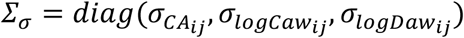, i.e., 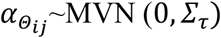.

Level 3: Participant

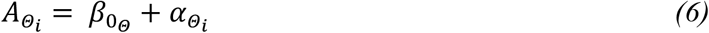

In the third level,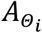 is decomposed into 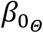, the overall population-mean NO parameters when *X*_*ij*_ =0 and the otherwise unexplained between-participant variation in the NO parameters, represented by the participant-level random intercepts 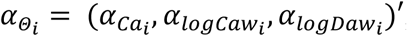, assumed to follow a MVN with variance-covariance matrix *Σ* _*τ*_, i.e., 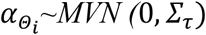.

Levels 2 and 3 are combined in the following equation with two random intercepts: one at the visit level and the other at the participant level.

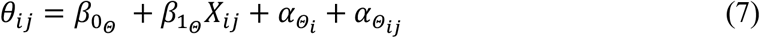

Prior distributions for L-U-HB are specified to be relatively non-informative. We assume the regression coefficients each have independent multivariate normal priors (with ***I*** indicate a square identity matrix):

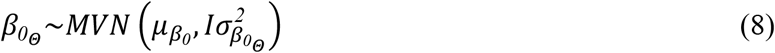

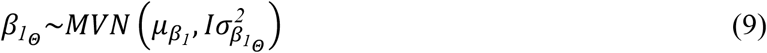

with 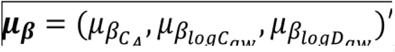, a non-informative prior distribution using large variances 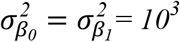. The random intercept variance-covariance matrix is assumed to have a non-informative inverse-Wishart prior distribution with non-informative diagonal matrix D=diag (0.001, 0.001, 0.001). And the variances of within-participants are sampled from non-informative independent inverse gamma distributions. Finally, the residual variance *σ*^*2*^ is assumed to have a non-informative inverse-Gamma distribution, *Inv-Gamma* (0.001, 0.001).

The L_U_HB and L_TS_HB’s first stage was simulated via JAGS as mentioned above. The simulation process includes an adaptive mode phase (“burn-in”) and a long enough updating phase [25]where the adaptive mode is turned off.

#### 6. L-U-NLME

The longitudinal version of the unified NLME model is similar to the cross-sectional one in which the covariate is linked to the mean function for NO parameters, except that it also specifies the variance-covariance matrix for the visit level subgroup. We also specify the diagonal matrix for the visit level variations for our simplified model.

### Constraint on C_A_

C_A_ must be non-negative since it represents the concentration, in ppb, of the NO in the alveolar compartment [12]. There are similar constraint considerations for C_aw_ and D_aw_ which we satisfy by modeling logC_aw_ and logD_aw_ since C_aw_ and C_aw_ to have approximately log-normal population-level distributions. C_A_ tends to have more of a normal or truncated normal distribution, so a different approach is necessary. In the simulation study data generation step (described below), we discard samples if their C_A_ was negative. We also enforce the non-negative constraint in the HB models by using a truncated distribution function. We didn’t apply the constraint on HMA since our previous papers [10, 12] proved that it had poor performance, probably due to the large number of failed to converge in Stage I. The NLME models implemented in the nlme package in R were fitted without such constraints since there are no readily available constraint options. Constrained versions of TS_NLS proved to be more biased in our previous study[12], thus we implement only the unconstrained NLS here.

### Simulation study

We compare the above methods in an extensive simulation study, roughly based on the CHS study design. Each simulated dataset consists of 500 participants with 3 visits each and each visit includes 8 multiple flow FeNO maneuvers (2 each at: 30, 50, 100, and 300 ml/s), which we simulate under a given “scenario” of underlying true associations of the NO parameters with a standard normal covariate X_ij_ (independent across and within participants). For a given scenario, 100 replicate datasets (each of N* M =500*3) are generated. Data-generating values, shown in Table 1, of population-level mean NO parameters 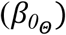, the between-participant variance-covariance matrix 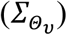, and the residual variance 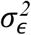 are based on values estimated in a preliminary L-TS-NLME analysis of CHS data and are similar to the values in the previous cross-sectional study[12]. We set the with-participant covariances to be zero for simplicity.

**Table 1.** Parameter values used to generate data in the simulation study

| NO parameter | Population-level (across-participant) |  | Within-participant (across-visit) | Population-level (across-participant) |  |
| --- | --- | --- | --- | --- | --- |
| | Mean ($\mu$) | Standard Deviation ($\tau$) | Standard Deviation ($\sigma$) | Correlation ($\rho$) | |
| $C_A$ | 1.5 | 0.45 | 0.50 | $C_A, \log_{Caw}$ | 0.66 |
| $\log_{Caw}$ | 3.5 | 0.65 | 0.40 | $C_A, \log_{Daw}$ | -0.38 |
| $\log_{Daw}$ | 2.5 | 0.55 | 0.33 | $\log_{Caw}, \log_{Daw}$ | -0.35 |

The different scenarios for the simulation study are described in Table 2. The regression coefficients 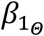 relating X_ij_ to each NO parameter take a range of values: 0.01, 0.05 to 0.1. In Scenario 1, the reference scenario, all NO parameters have the same association with the covariate 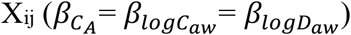, and the value of this association is either 0.01, 0.05 or 0.1.

**Table 2.**
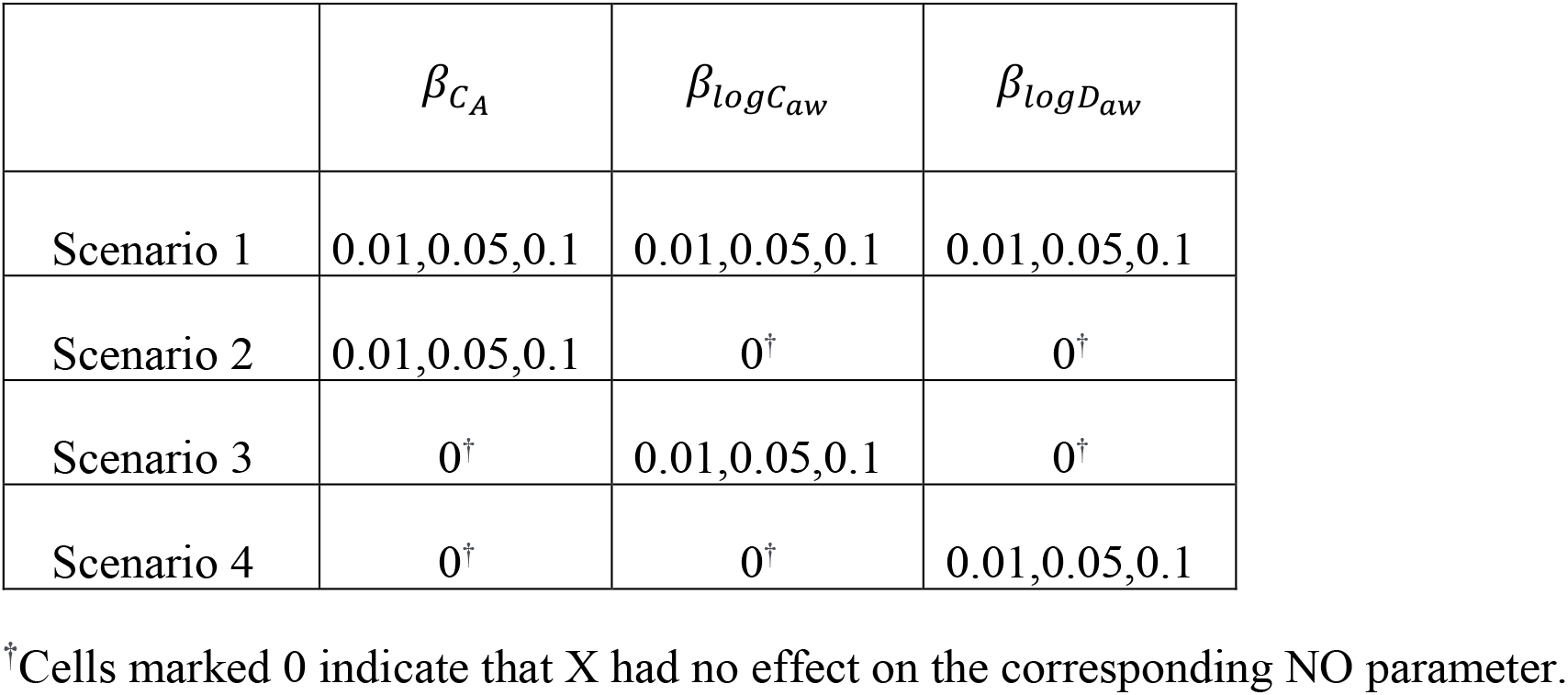
The 4 simulation study scenarios are each repeated at 3 effect sizes and replicated 100 times, for 1200 simulated datasets in total.

In this simulation study we compare performance of the methods based on several metrics: percent bias (% bias), 95% Confidence/Credible (CI) length and coverage, power and type I. Percent bias is calculated as: (Estimate-True value)/True value for non-zero parameters. The 95% CI length and coverage are calculated using the given 95% confidence interval for frequentist approaches and 95% credible interval (using the returned posterior distribution, the center portion contains 95% of the values) for Bayesian approaches. In primary analyses, Scenario 1 is used to calculate bias, power, 95% CI coverage, 95% CI length and the other scenarios (S2-S4) are used to calculate type I errors. In secondary analyses, Scenarios 2-4 are used to calculate bias, power & type I error rate, 95% CI coverage, 95% CI length.

### CHS data analysis

We analyzed data from a CHS cohort originally recruited in kindergarten/1^st^ grade [26] with FeNO50 assessed at 6 study visits over 8 years (spanning ages 8-16) and multiple flow FeNO assessed at the last 2 study visits, when most children were ages 13-14 and 15-16. The CHS multiple flow FeNO protocol called for 9 maneuvers at each of four expiratory flow rates (3 at 50 ml/s and 2 each at: 30, 100, and 300 ml/s) collected using chemiluminescence analyzers (model CLD88-SP with DeNOx accessory to provide NO-scrubbed air; EcoMedics, Duernten, Switzerland/Ann Arbor, MI, USA) as described in detail elsewhere [27, 28]. FeNO data processing was based on the ATS/ERS guidelines for FeNO at 50 mL · s−1 [7] with a search window based on airway turnover[29]. Each CHS child participant provided informed assent and a parent/guardian provided informed consent. The CHS data were collected using a protocol approved by the University of Southern California Institutional Review Board, the analyses in this paper were conducted under HS-13–00150, and all methods were carried out in accordance with relevant guidelines and regulations.

In a previous longitudinal analysis, FeNO_50_ was found to have a strong positive linear association with height across this age range in children without asthma [30]. To complement the previous analysis relating longitudinally assessed FeNO_50_ to height, here we relate longitudinally assessed NO parameters (from the up to 2 repeated assessments of multiple flow FeNO) to standardized height (population-mean centered: 162.7 centimeters and population-SD scaled: 8.75 centimeters). The analyses included 1004 children who never reported a doctor diagnosis of asthma and had multiple flow FeNO data available at both visits. The average number of valid multiple flow maneuvers at the first and second visits were 9.68 and 8.97, respectively.

## Results

### Simulation study

Computation time was longer for unified methods than for two-stage methods, as expected. The computation time for a given method was similar across scenarios and slightly shorter for larger β coefficients (Supplementary Table 2). Average computation times on a high-performance computing platform (3 CPU, 12GB memory) for a single simulated dataset (500 participants, 3 visits each, 8 maneuvers per visit) were: 30 hours for L_U_HB, 23 hours for L_TS_HB, 14.7 minutes for L_U_NLME, 11.4 minutes for L_TS_NLME, 4.6 seconds for L_TS_HMA and 6.2 seconds for L_TS_NLS. Most methods had reasonable convergence rates (99% for L_TS_HB, 93% for L_U_HB, 94% for L_TS_NLME) except for L_U_NLME (51%). L_TS_NLS and L_TS_HMA converged for all datasets, however Stage I of L_TS_NLS had 29% of participant models fail to converge on average (resulting in the exclusion of these participants’ results in Stage II) while L_TS_HMA had only 0.016% failures in Stage I (Supplementary Figure 2). The following simulation study results are a summary of all available converged results, since the intersection of datasets which converged under all methods is relatively small due to the convergence issue for L_U_NLME.

Figure 2 compares the percent bias and 95% CI interval properties for estimation of *β*_*Ca*_, *β*_*logCaw*_, *β*_*logDaw*_, across methods. For many methods, percent bias tended to decrease as the effect size of the covariate increased. Among all methods, L_U_HB had the lowest absolute values of percent bias for all three NO parameter associations (all < 4%), however at a small magnitude effect size (true β of 0.01) there was 52% bias for *β*_*logDaw*_, equivalent to a 0.052 bias on the original scale. L_U_NLME also had good performance in the subset of datasets where the method converged. Two stage methods (L_TS_HB, L_TS_NLME, L_TS_NLS) tended to have negative percent bias for *β*_*Ca*_, *β*_*logDaw*_ and positive percent bias for *β*_*logCaw*_. For a given method and effect size, percent bias was smaller for *β*_*Ca*_ than for associations with other NO parameters, perhaps because C_A_ is in the linear part of the 2CM for FeNO. In simulation Scenario 3, when only logC_aw_ had an association with X, the directions of bias for two stage methods were different from other scenarios (Supplemental Figure 1.C1). But L_U_HB and L_U_NLME still had the smallest bias. An alternative version of L_TS_HMA with the constraint that Stage I C_A_>0 in Scenario 1 (Supplementary Figure 3) resulted in ~30% of Stage I estimates being dropped, on average, and patterns in percent bias similar to other two stage methods.

**Figure 2:**
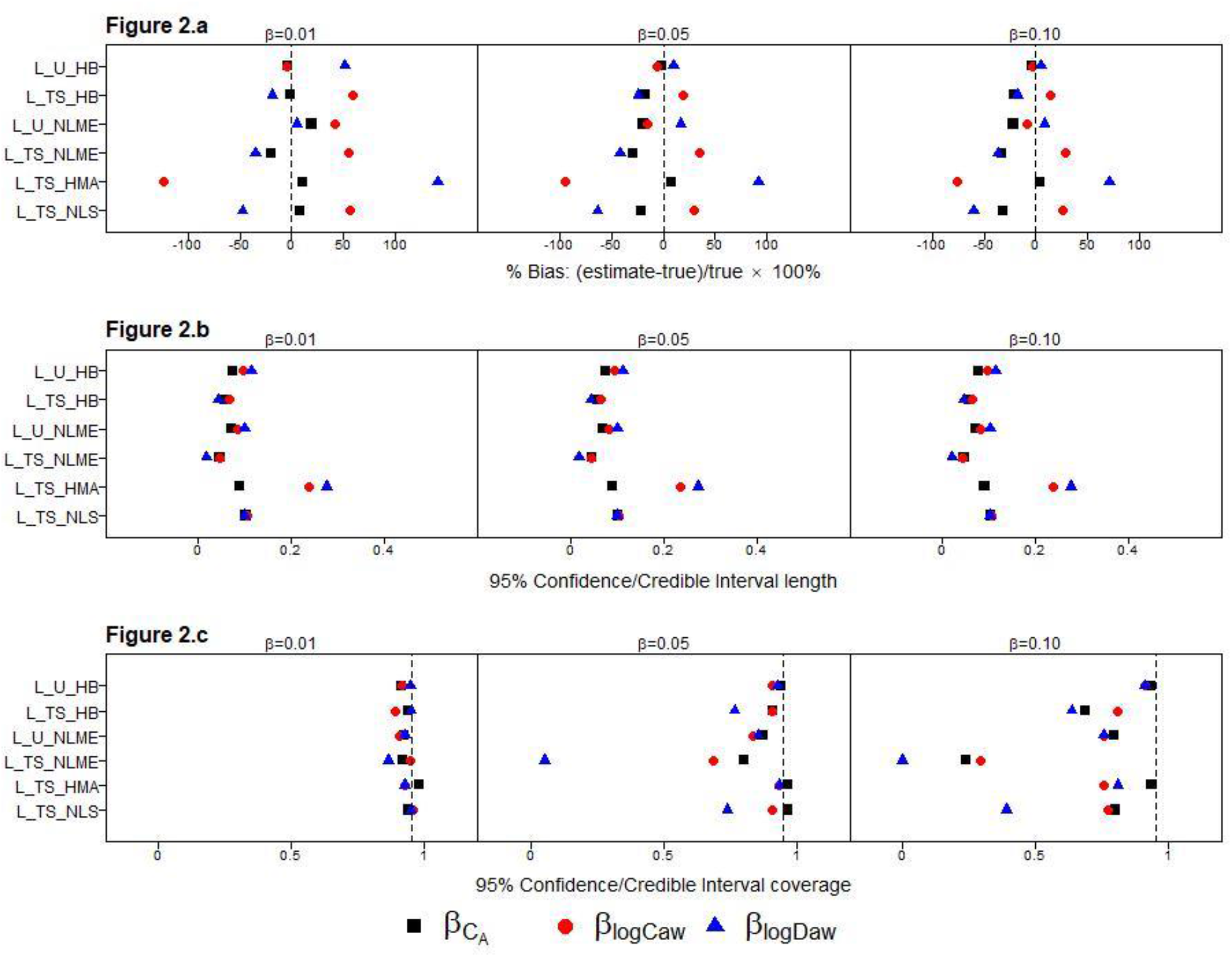
Comparison of method performance in terms of: percent bias (a), 95% CI length (b), and 95% CI coverage (c) for estimating associations with NO parameters (*β*_*Ca*_: black square, *β*_*logCaw*_: red circle, *β*_*logDaw*_: blue triangle) in simulation study Scenario 1, replicated at 3 different effect sizes (*β*= 0.01, 0.05 or 0.1).

For a given method, the 95% CI lengths (Figure 2b) were similar across NO parameters and these patterns did not vary based on magnitude of the true β. L_TS_NLME had the shortest 95% CI lengths followed by L_TS_HB. For 95% CI coverage (Figure 2c), coverage declined as the magnitude of the true β increased, for all methods except L_U_HB. It was the only method to produce 95% CIs with approximately appropriate coverage (91 ~ 95%) for all s. L_U_NLME had slightly larger biases and shorter 95% CI lengths, resulting in 75 ~ 80% coverage when the true β was 0.1. The higher coverage for these two unified methods was due to a combination of low bias and longer 95% CI. L_TS_HB also had reasonable coverage, but it differed across NO parameters, ranging from 63 ~ 81%. For L_TS_NLME, L_TS_HMA, and L_TS_NLS, low coverage was due to a combination of large bias and short CI lengths.

Power curves and Type I error rates for *β*_*Ca*_, *β*_*logCaw*_, *β*_*logDaw*_ across all simulation scenarios are shown in Supplementary Figure 1. For simplicity, here Figure 3 displays a subset of these data: the power for each NO parameter association versus two versions of type I error, based on scenarios 2-4 at the largest magnitude effect size considered (true β of 0.1). Ideally, a method will produce 2 values in the upper left-hand corner of this plot, which indicates high power and low type I error regardless of which another NO parameter had a non-zero association. Indeed, L_U_HB had relatively high power (1.00 for *β*_*Ca*_, 0.97 for *β*_*logCaw*_, and 0.89 for *β*_*logDaw*_) and low type I error rates from 0.02 to 0.07 for all three NO parameter associations except 0.12 for *β*_*logCaw*_ in Scenario 4 where only *β*_*logDaw*_ was non-zero. Other methods generally also had good power, but L_TS_NLS had low power for *β*_*logDaw*_ and L_TS_HMA had low power for both *β*_*logCaw*_ and *β*_*logDaw*_. Except for L_U_HB, most methods had inflated type I error for *β*_*logCaw*_ when *β*_*logDaw*_ was non-zero, or vice versa. For example, L_TS_HB had excellent power for *β*_*Ca*_ (0.98) and low type I error (0.02) when *β*_*logCaw*_ was nonzero but higher type I error (0.17) when *β*_*logDaw*_ was nonzero. This issue became more pronounced for *β*_*logCaw*_, where L_TS_HB again had excellent power for *β*_*logCaw*_ (1.00) and low type I error (0.01) when *β*_*Ca*_ had a nonzero association but very high type I error (0.69) when *β*_*logDaw*_ was nonzero. In summary, L_U_HB had high power and low type I error rates for all NO parameter associations while other methods had good power and type I error for *β*_*Ca*_ and good power but inflated type I error rates for *β*_*logCaw*_ and *β*_*logDaw*_. Exceptions to this pattern were L_TS_NLS and L_TS_HMA, both of which had low power for *β*_*logDaw*_ due to their large negative biases.

**Figure 3:**
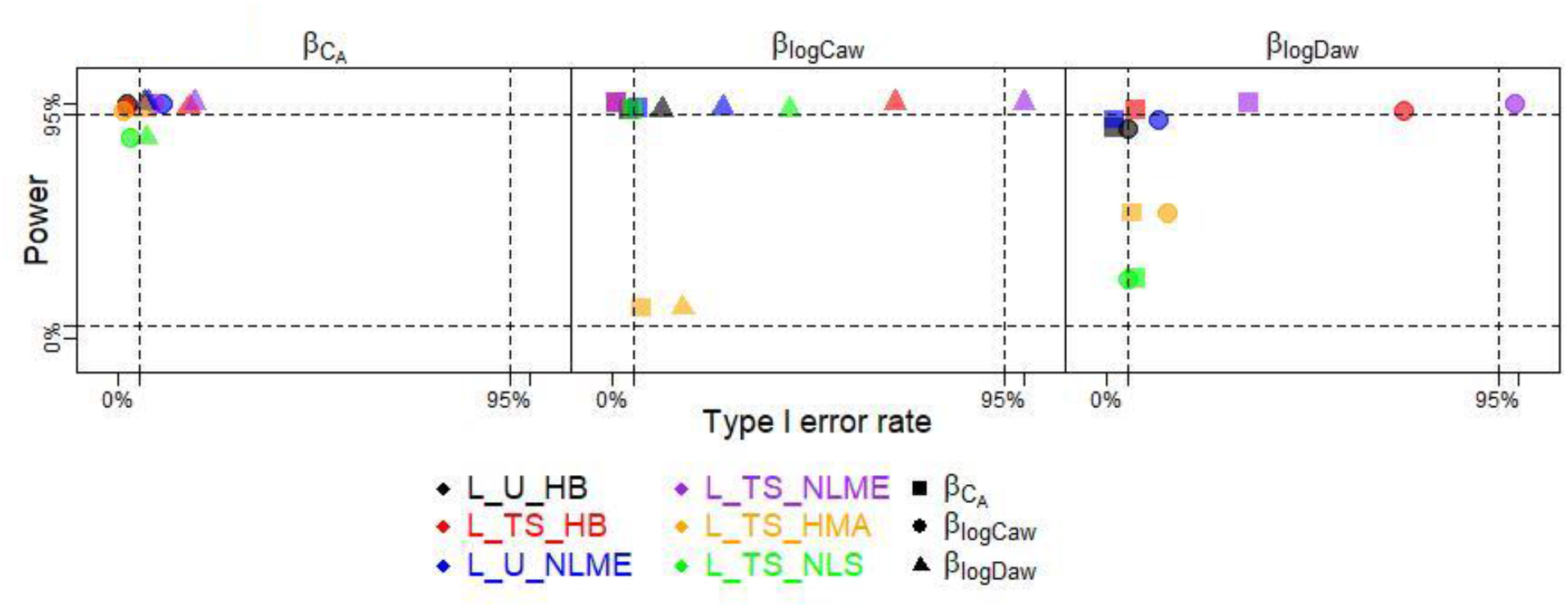
Power and Type I errors for the 6 methods (distinguished by color) for true β of 0.1, with power for a given NO parameter’s association calculated from the simulation scenario where only that association is non-zero (e.g., power for *β*_*Ca*_ from Scenario 2 where *β*_*Ca*_ = 0.1, *β*_*logCaw*_ = 0, *β*_*logDaw*_ =0) and Type I error calculated under two scenarios (e.g., Type I error for *β*_*Ca*_ under S3: *β*_*Ca*_ =0, *β*_*logCaw*_ =0.1 *β*_*logDaw*_ = 0 and under S4: *β*_*Ca*_ =0, *β*_*logCaw*_ =0, *β*_*logDaw*_ =0.1), with the non-zero NO parameter association denoted by shape.

While the primary focus of the simulation study was on the estimation of *β*_*Ca*_, *β*_*logCaw*_, and *β*_*logDaw*_, we also studied estimation of random effect variances at the participant and visit levels. Participant-level variances, particularly for C_A_ and logD_aw_, tended to be underestimated by most methods, though L-U-HB had the overall lowest bias (Supplemental Figure 1) Participant-level correlations also tended to be underestimated (Supplemental Figure 1).

In summary, this simulation study demonstrated that L_U_HB generally had the smallest bias, appropriate 95% CI coverage, and good power with low type I error rates for all three NO parameter associations across scenarios. L_TS_HB, the two-step version of L_HB, had greatly reduced computation time and maintained good power at the expense of introducing some bias, poorer 95% CI coverage at larger magnitude effects, and high type I errors for *β*_*logCaw*_ and *β*_*logDaw*_. Compared to L_TS_HB, L_U_NLME had similar performance, and even less inflated type I error rates but failed to converge in ~50% of the simulated datasets. Compared to L_TS_HB, L_TS_NLME had bias in the same direction but of larger magnitude, resulting in lower coverage and more inflated type I errors. L_TS_NLS, on average, had ~40% of participants fail to have Stage I estimates (Supplementary Table 1). Despite this, L_TS_NLS performed well for estimation of *β*_*CA*_, but for *β*_*logCaw*_ and *β*_*logDaw*_ had considerable bias, low power and inflated type I error.

### CHS data analysis

Applying the 6 methods to a CHS analysis relating NO parameters to height, we observed associations with height that were: positive or null for C_A_, positive for logC_aw_, and negative or null for logD_aw_ (Figure 4). The two unified methods (L_U_HB and L_U_NLME) both estimated similar statistically significant associations between height and all three NO parameters.

**Figure 4:**
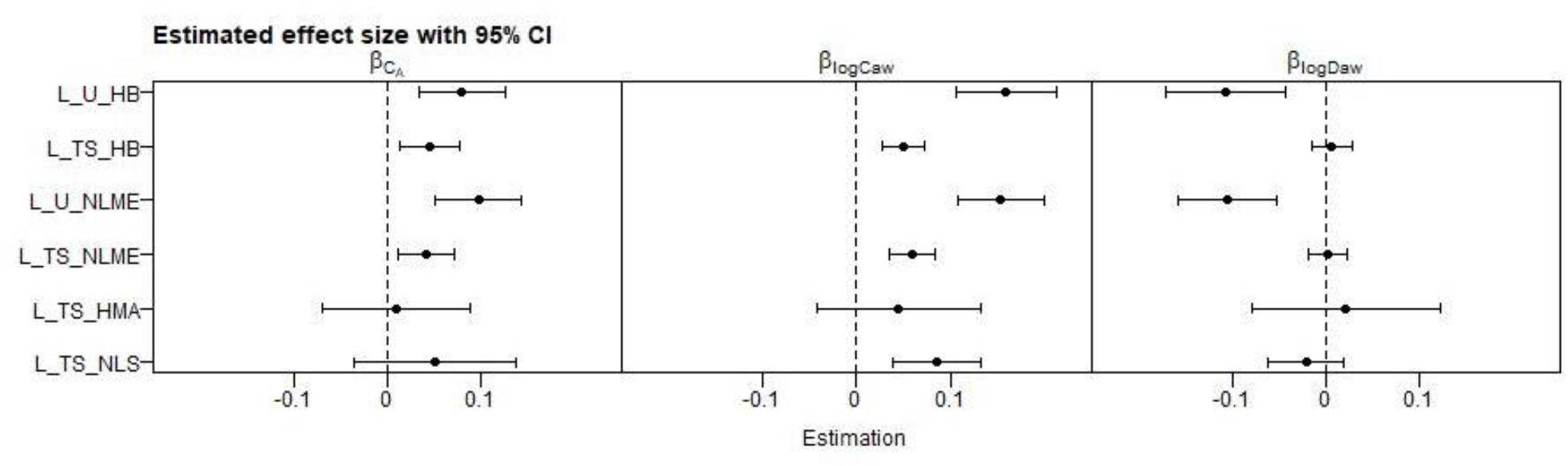
Estimated associations between NO parameters and standardized height in the CHS using 6 methods.

Specifically, from the L_U_HB model we estimated that, a within-participant increase in height of 8.79 centimeters was associated, on average, with a 0.079 (95% CI: 0.034, 0.125) ppb increase in C_A_, a 0.158 (95% CI: 0.106, 0.212) increase in logC_aw_, and a 0.106 (95% CI: 0.044, 0.171) decrease in logD_aw_. These latter two estimates are equivalent to a 17% increase in C_aw_ and a 10% decrease in D_aw_. From the L_U_NLME model, analogous estimates were similar: 0.092 (95% CI: 0.046, 0.138) for C_A_, 0.149 (95% CI: 0.104, 0.194) for logC_aw_, -0.104 (95% CI: -0.157, -0.052) for logD_aw_). Two-stage methods produced lower estimates for *β*_*Ca*_ and *β*_*logCaw*_, and higher estimates for *β*_*logDaw*_. Furthermore, two-stage method estimates were all approximately null and non-significant for *β*_*logDaw*_. Estimates of *β*_*Ca*_ were not statistically significant for L_TS_NLS and L_TS_HMA. There was a clear pattern when comparing a unified method to its two-stage counterpart (i.e., L_U_HB vs L_TS_HB and L_U_NLME vs L_TS_NLME) in that the unified method had larger magnitude estimates (farther from zero) than their corresponding two-stage versions. The finding that the unified methods produced more significant estimates than two-stage ones (especially the L_TS_NLS and L_TS_HMA), which agreed with results in our simulation. For L_TS_NLS, a Stage I outlier (−1325) for C_A_ resulted in an extremely wide 95% CI for *β*_*Ca*_ in Stage II, so we excluded this outlier for Figure 4. Stage I convergence failures were observed for 528 and 30 out of 1004 models for L_TS_NLS and L_TS_HMA, respectively.

## Discussion

In this paper, we proposed a novel unified hierarchical Bayesian model, L_U_HB, for relating longitudinally assessed NO parameters to covariates, extending our previous cross-sectional unified hierarchical Bayesian model, and performed the first evaluation of the statistical properties of various two-stage methods for longitudinal analysis of NO parameters. In a simulation study, L_U_HB performed well for estimating associations of NO parameters with covariates, with small bias, appropriate 95% CI coverage, good power, and low type I error rates. The two-step version, L_TS_HB, had greatly reduced computation time and maintained good power at the expense of introducing some bias, poorer 95% CI coverage at larger magnitude effects, and high type I errors for *β*_*logCaw*_ and *β*_*logDaw*_. The other unified method L_U_NLME had good performance when it converged, but it had serious convergence issues. Other two-stage methods had drawbacks in terms of bias, inflated type I error rates, etc.

In a previous simulation study comparing the performance of methods estimating NO parameter associations with a covariate in a cross-sectional study [12], U_HB had the best performance across all simulation scenarios, similar to our findings in this longitudinal study. L_U_NLME had the second-best performance across all three NO parameters in the longitudinal study, while its cross-sectional version (U_NLME) had large bias in estimating *β*_*Ca*_. L_TS_NLS also had much better performance in estimating *β*_*Ca*_ and *β*_*logCaw*_ in the longitudinal study than TS_NLS for the cross-sectional study but still had large bias for *β*_*logDaw*_. Both L_TS_NLME and the cross-sectional TS_NLME had large bias and poor coverage.

Computation time is an issue. L_U_HB had superior statistical properties to many competitor methods, albeit at additional computational expense. The cross-sectional U_HB model had an average computation time of 5.5 hours for N=1000 participants (ref) while for the longitudinal version (L_U-HB) average computation time was 30 hours for N=500 participants, 3 visits each. While the longer L_U_HB computation time was burdensome in a simulation study with 1000s of datasets, it is less of an issue when analyzing a single dataset. However, note that the computational cost will increase as more covariates are added, or a more complex model is used in Stage II. The other competitive method was L_U_NLME, which had the second-best estimation performance and ran faster, but it had poor convergence. Our results suggest that for further applications in which there are more variables or more complex models, where the unified model may become computational intractable, a two-stage version of hierarchical Bayesian model will have reasonably good performance and therefore be used in iterative model building, with a single run of L_U_HB for the final results, using L_TS_HB estimates as starting values to speed convergence.

For biologically plausibility, we constrained *C*_A_ to be non-negative. In our previous cross-sectional study, we encountered a problem constraining C_A_ to be non-negative in the standard JAGs software. We solved it by sampling logC_aw_ and logD_aw_ as they were bivariate normal distributed, and then sample zero-truncated C_A_ conditioned on them. In that case, we sampled the variances and correlations one by one and set boundaries for the last correlation to ensure a 3×3 positive definite variance covariance matrix. The same solution was also used for L_U_HB and L_TS_HB when we set NO parameters to be correlated in both levels. But in our simplified model which assumed no correlations in the visit level, the randomness and constraint on C_A_ were easily specified separately without considering the validity of the variance-covariance matrix.

When applying L_U_HB to study the association between height and NO parameters using longitudinal multiple flow FeNO data on healthy schoolchildren in the CHS, we found positive associations of C_A_ and logC_aw_ with height and a negative association of logD_aw_ with height. Had we applied only L_TS_HMA or L_TS-NLS methods, we would have failed to detect associations of height with C_A_ or logD_aw_. Our findings add to the limited literature on associations of NO parameters with height/age. A previous analysis using longitudinal FeNO_50_ data from the same cohort over a longer follow-up period, from ages 8-16, found that FeNO_50_ increased approximately linearly with height and FeNO_50_ increased nonlinearly with age [30].

Limited cross-sectional data on trends in NO parameters by age (for participants less than 20 years old) suggests non-significant increases of D_aw_ and C_aw_ but a decrease in C_A_ though some influential values for the oldest participants in the sample may have impacted these results [31]. Additionally, differences with our findings may be due to a different age range or due to the difference between a cross-sectional (between-person) vs longitudinal (within-person) design.

There are several directions for future work. To reduce computation time observed when implementing L_U_HB with JAGS, alternative GIBBS sampling software such as Rstan could be considered. Several components of the model implementation appeared to affect convergence rates, such as the number of parameters or the length of adaptation phase. The length of adaptation was the most important factor, but adaptation is in itself a computationally intensive operation. Given the computational costs involved, we chose to base our simulation study on smaller dataset sizes (500 participants, 3 visits each) and simplify our model so that it could converge within two days on average. The simplified version of our longitudinal model ignored the correlation between the NO parameters within the participant level (V0 model). L_U_HB and L_TS/U_NLME were able to specify the diagonal matrix for visit-level variation. While the NLS and the HMA approaches fitted the FeNO models for each observation, thus ignoring any correlations between or within participants. In the CHS data analysis, we only selected participants who have visited both in year 8 and 10 as a more direct comparison to the simulation study, but unified models are capable of handling unbalanced data.

In conclusion, in this paper we presented a longitudinal extension of the unified hierarchical Bayesian model for analyzing nonlinear data (e.g., FeNO data). Despite the long computation time required for achieving convergence, L_U_HB had the best performance estimating covariate coefficients as well as variance-covariance components. Its two-stage version L_TS_HB served as a decent alternative to obtain a faster pilot estimation for much more complicated models.

## Supporting information

no link

## Data Availability

Due to limitations in the orignal consent forms and HIPAA requirements, data from the CHS cannot be freely available in the public domain. However, we are committed to sharing the data and results acquired as part of this study. The CHS has a process in place for data sharing that involves approval of proposals by a Data Sharing Committee. Investigators who want access to data will be required to submit a research protocol, which will be reviewed by the CHS Health Data Release Committee and the USC IRB. Recipients must agree to security policies. Please send requests to access this dataset to Dr. Sandrah Eckel.

## Data availability

Due to limitations in the original consent forms and HIPAA requirements, data from the CHS cannot be freely available in the public domain. However, we are committed to sharing the data and results acquired as part of this study. The CHS has a process in place for data sharing that involves approval of proposals by a Data Sharing Committee. Investigators who want access to data will be required to submit a research protocol, which will be reviewed by the CHS Health Data Release Committee and the USC IRB. Recipients must agree to security policies. Please send requests to access this dataset to Dr. Sandrah Eckel.

