## Supplementary material for "Longitudinal Hierarchical Bayesian models of covariate effects on airway and alveolar nitric oxide": no link

Supplementary materials

We now show a number of further details of results of our simulation study. We begin by exploring convergence rates across a range of methods and scenarios.

### Table 1a：Number of simulation study datasets (out of 100) with successful convergence for each method and scenario.

| Method | Scenario 1 | | | Scenario 2 | | | Scenario 3 | | | Scenario 4 | | |
| --- | --- | --- | --- | --- | --- | --- | --- | --- | --- | --- | --- | --- |
|  | $\beta_{Ca}$ = $\beta_{logCaw}$ = $\beta_{logDaw}$ | | | $\beta_{logCaw}$ = $\beta_{logDaw}$ = 0 | | | $\beta_{Ca}$ = $\beta_{logDaw}$ = 0 | | | $\beta_{Ca}$ = $\beta_{logCaw}$ = 0 | | |
|  |  |  |  | $\beta_{Ca}$ | | | $\beta_{logCaw}$ | | | $\beta_{logDaw}$ | | |
|  | 0.01 | 0.05 | 0.1 | 0.01 | 0.05 | 0.10 | 0.01 | 0.05 | 0.10 | 0.01 | 0.05 | 0.10 |
| L_U_HB | 95 | 100 | 89 | 89 | 97 | 98 | 91 | 88 | 95 | 90 | 97 | 92 |
| L_TS_HB | 100 | 99 | 99 | 97 | 99 | 100 | 98 | 100 | 100 | 100 | 100 | 99 |
| L_U_NLME | 54 | 55 | 49 | 49 | 48 | 48 | 46 | 45 | 47 | 56 | 55 | 56 |
| L_TS_NLME | 96 | 96 | 93 | 96 | 95 | 93 | 95 | 96 | 92 | 93 | 92 | 96 |
| L_TS_HMA | 100 | 100 | 100 | 100 | 99 | 100 | 100 | 100 | 100 | 100 | 100 | 99 |
| L_TS_NLS | 100 | 100 | 100 | 100 | 100 | 100 | 100 | 100 | 100 | 100 | 100 | 100 |
| All methods converged | 53 | 54 | 41 | 45 | 45 | 47 | 41 | 39 | 44 | 51 | 54 | 50 |

**Table 1b：Average Stage I convergence failure rates* for select two-stage methods: L_TS_HMA and L_TS_NLS.** ^[1]^

| Method | Scenario 1 | | | Scenario 2 | | | Scenario 3 | | | Scenario 4 | | |
| --- | --- | --- | --- | --- | --- | --- | --- | --- | --- | --- | --- | --- |
|  | $\beta_{Ca}$ = $\beta_{logCaw}$ = $\beta_{logDaw}$ | | | $\beta_{logCaw}$ = $\beta_{logDaw}$ = 0 | | | $\beta_{Ca}$ = $\beta_{logDaw}$ = 0 | | | $\beta_{Ca}$ = $\beta_{logCaw}$ = 0 | | |
|  |  |  |  | $\beta_{Ca}$ | | | $\beta_{logCaw}$ | | | $\beta_{logDaw}$ | | |
|  | 0.01 | 0.05 | 0.1 | 0.01 | 0.05 | 0.10 | 0.01 | 0.05 | 0.10 | 0.01 | 0.05 | 0.10 |
| L_TS_HMA | 0.2 | 0.2 | 0.3 | 0.2 | 0.2 | 0.3 | 0.3 | 0.3 | 0.2 | 0.3 | 0.3 | 0.2 |
| L_TS_NLS | 435.3 | 435.6 | 439.3 | 432.4 | 434.5 | 435.1 | 433.5 | 433.5 | 433.6 | 438.2 | 434.2 | 436.3 |

### Table 2: Average Computation time for each method and simulation scenario.

Average computation times on a high-performance computing platform (3 CPU, 8GB memory) for a single simulated dataset (500 participants, 3 visits each, 8 maneuvers per visit) were: 29 hours for L_U_HB, 24 hours for L_TS_HBL, 14.7 minutes for L_U_NLME, 11.6 minutes for L_TS_NLME, 4.7 seconds for L_TS_HMA and 6.1 seconds for L_TS_NLS. However, the computation times differed slightly across scenarios. Table 2 summarizes the computation time of each scenario for all methods.

| Scenario | Scenario 1 | | | Scenario 2 | | | Scenario 3 | | | Scenario 4 | | | overall |
| --- | --- | --- | --- | --- | --- | --- | --- | --- | --- | --- | --- | --- | --- |
|  | $\beta_{Ca}$ = $\beta_{logCaw}$ = $\beta_{logDaw}$ | | | $\beta_{logCaw}$ = $\beta_{logDaw}$ = 0 | | | $\beta_{Ca}$ = $\beta_{logDaw}$ = 0 | | | $\beta_{Ca}$ = $\beta_{logCaw}$ = 0 | | |  |
|  |  |  |  | $\beta_{Ca}$ | | | $\beta_{logCaw}$ | | | $\beta_{logDaw}$ | | |  |
|  | 0.01 | 0.05 | 0.1 | 0.01 | 0.05 | 0.10 | 0.01 | 0.05 | 0.10 | 0.01 | 0.05 | 0.10 |  |
| L_U_HB  (hours） | 28 | 32.3 | 28.4 | 33.4 | 31.1 | 31.5 | 31.6 | 30.2 | 30.2 | 30.4 | 28 | 30 |  |
| L_TS_HB  (hours) | 25.4 | 22.7 | 23.7 | 22.6 | 23 | 21.8 | 23 | 21.7 | 23.3 | 21.3 | 22.7 | 22.4 |  |
| L_U_NLME  (minutes) | 18.5 | 13.7 | 11.7 | 15.2 | 17.3 | 14.7 | 16.5 | 14.6 | 14.4 | 12.3 | 18.2 | 15.3 |  |
| L_TS_NLME  (minutes) | 12.4 | 11.4 | 11 | 12.2 | 12.1 | 12.6 | 12.7 | 11.1 | 11.9 | 12.9 | 13.4 | 12.5 |  |
| L_TS_HMA  (seconds) | 4.9 | 4.6 | 4.7 | 4.7 | 4.4 | 4.5 | 4.4 | 4.5 | 4.5 | 4.5 | 4.5 | 4.6 |  |
| L_TS_NLS  (seconds) | 6.1 | 6.2 | 5.8 | 6.2 | 5.8 | 5.9 | 5.7 | 5.9 | 6.1 | 5.8 | 6 | 5.8 |  |

### Figure 1: Extensive comparison of method performance in Simulation Scenarios with subplots for the different parameters

The performance of methods were compared in terms of bias, 95% CI length and 95% CI coverage for coefficients, populational means, participant level standard deviations, participant level correlations and visit level correlations of NO parameters ($\beta_{Ca}$: black square, $\beta_{logCaw}$: red circle, $\beta_{logDaw}$: blue triangle), replicated at 3 different effect sizes ($\beta$ = 0.01, 0.05 or 0.1). Power and type I error rates for coefficients. These results are shown below.

#### A: Scenario 1

| A1: Estimated Associations β |
| --- |
| 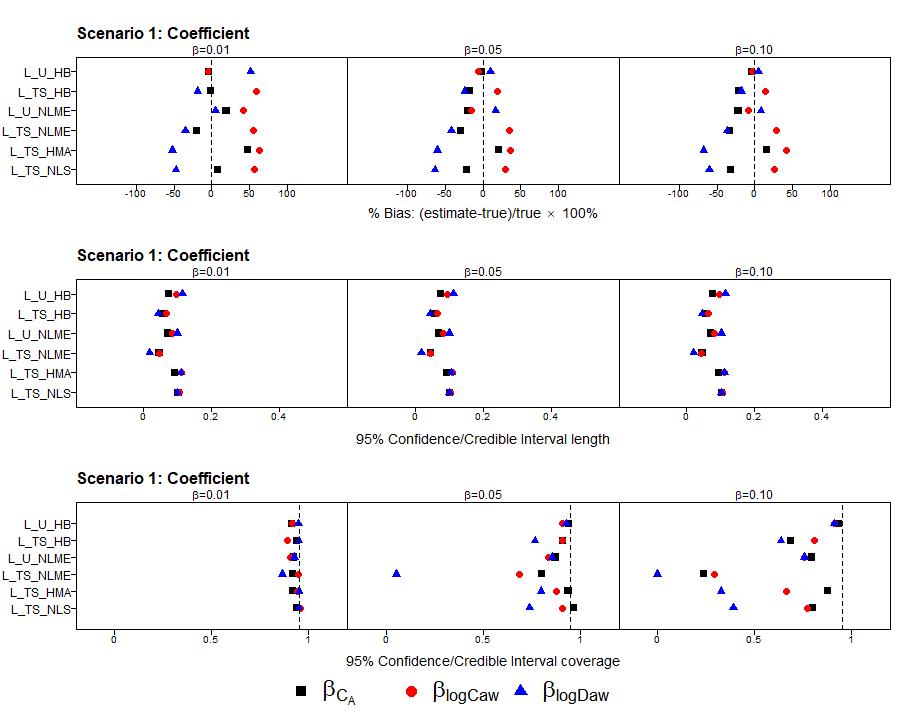 |
| 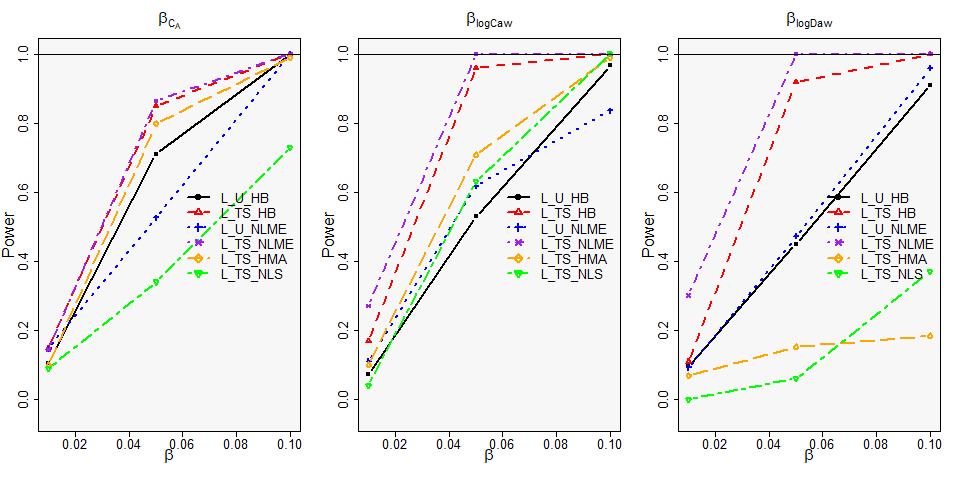 |

##### A2: Population level mean ɑ

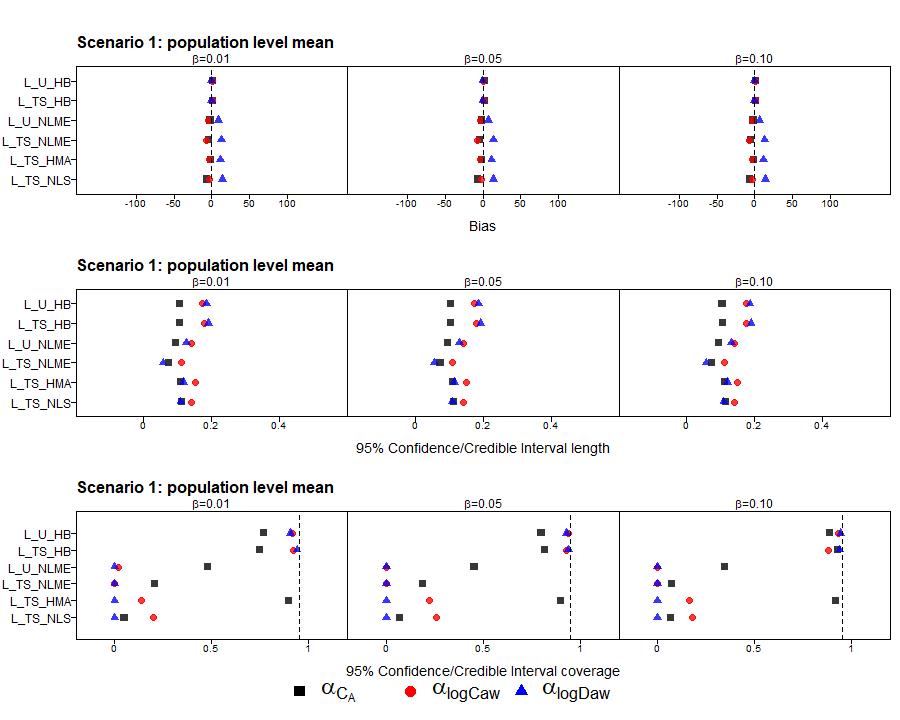

| A3: Participant level standard deviations τ |
| --- |
| 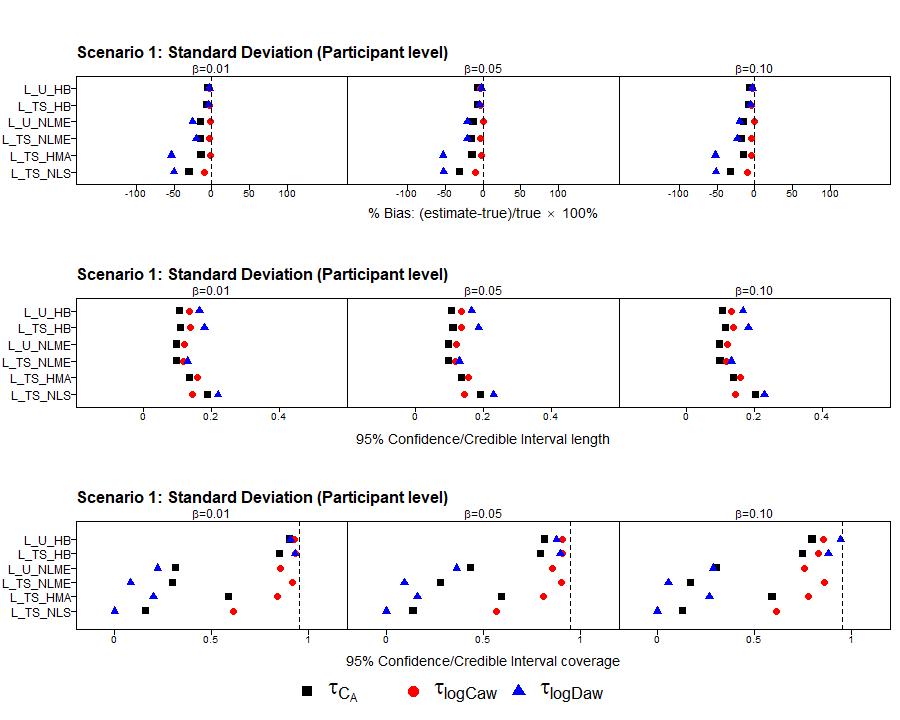 |

##### A4: Participant level correlations ⍴

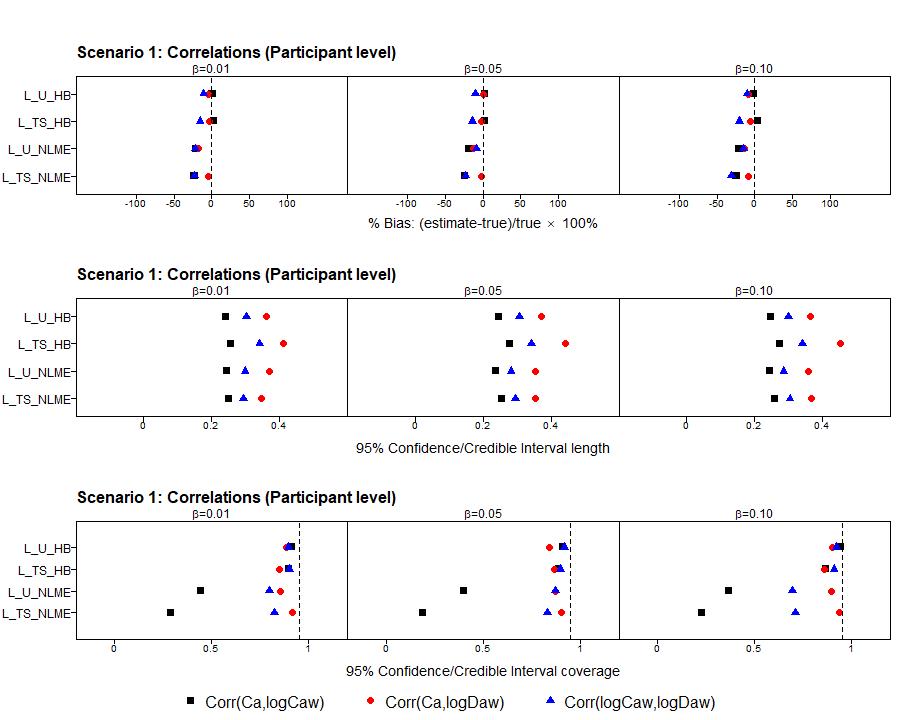

##### A5: Visit level standard deviations σ

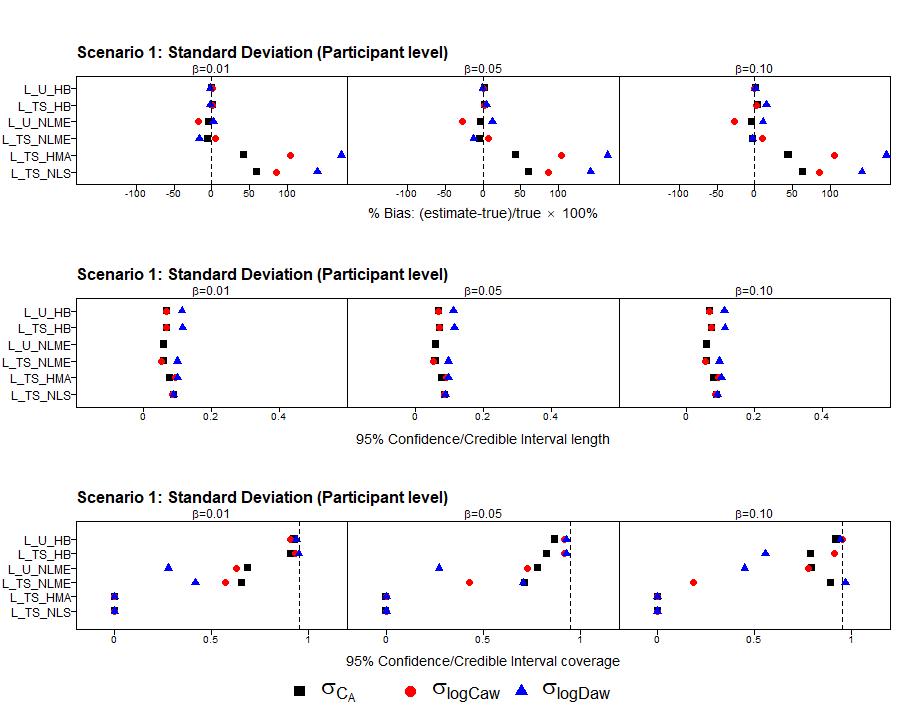

#### B: Scenario 2

| B1: Estimated Associations β (Bias instead of % Bias) |
| --- |
| 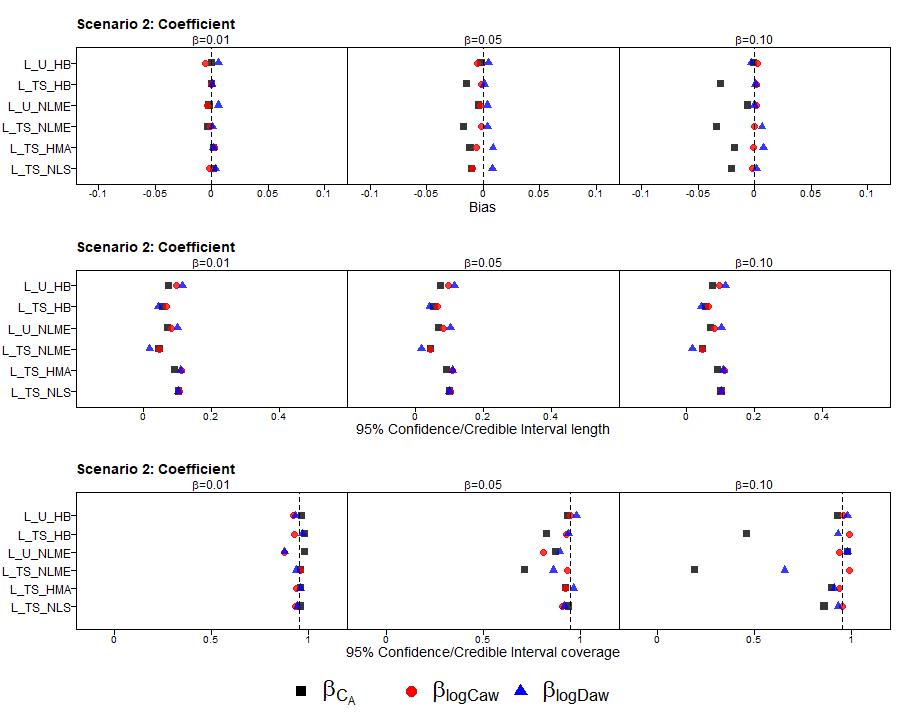 |
| 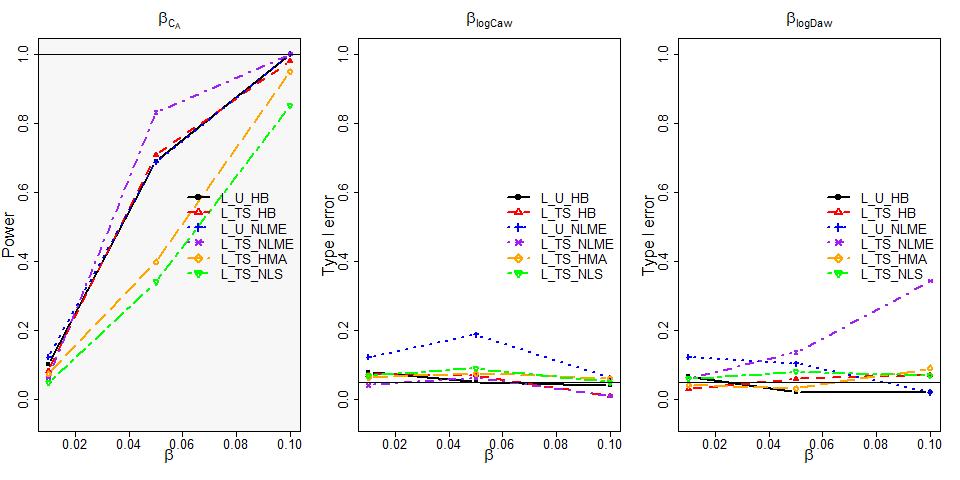 |

##### B2: Population level mean ɑ

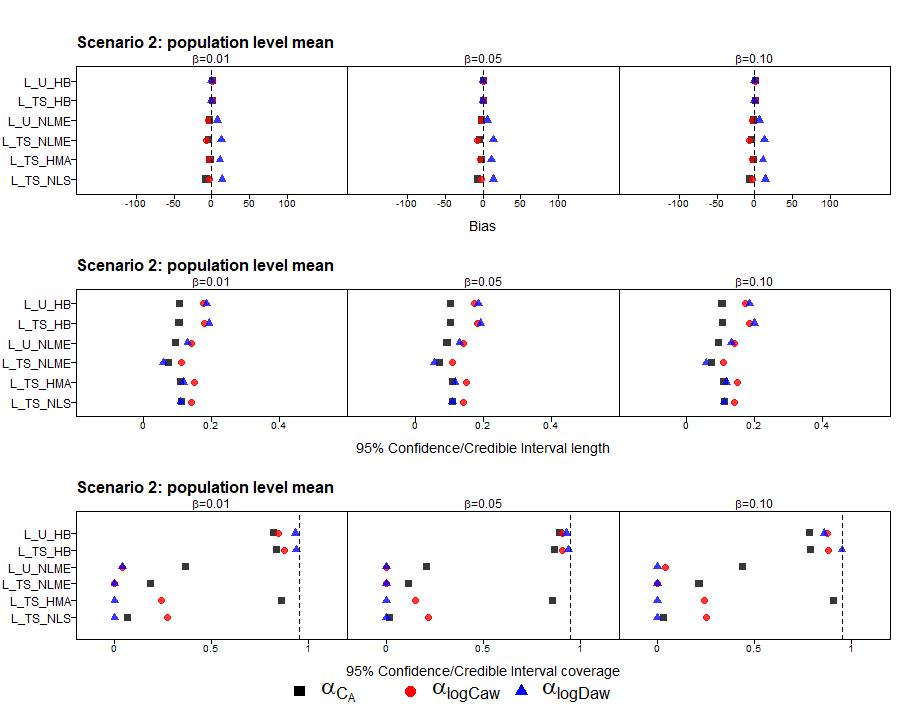

##### B3: Participant level standard deviations τ

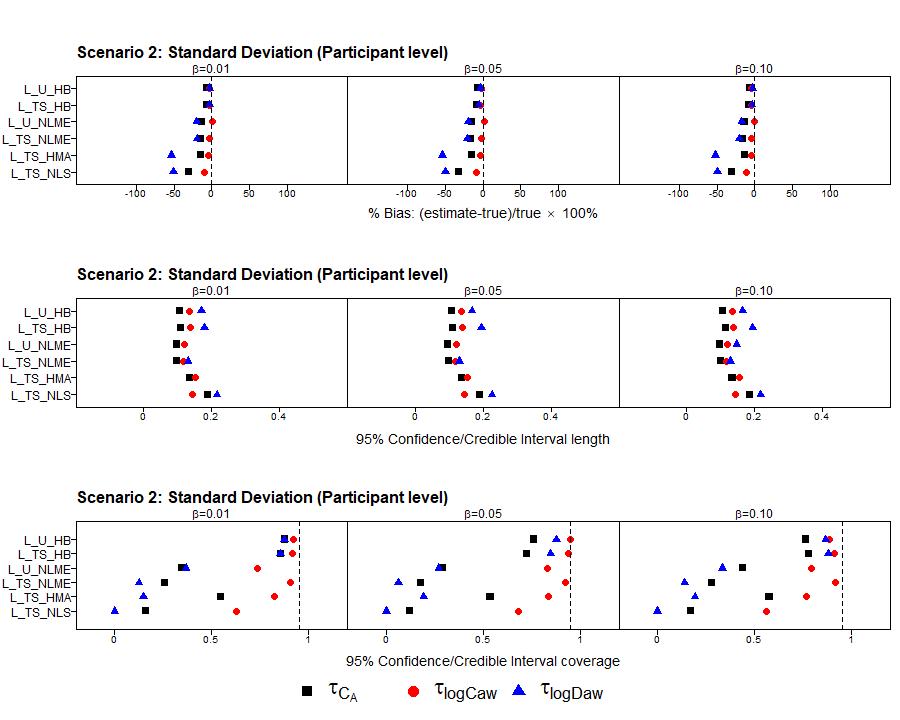

##### B4: Participant level correlations ⍴

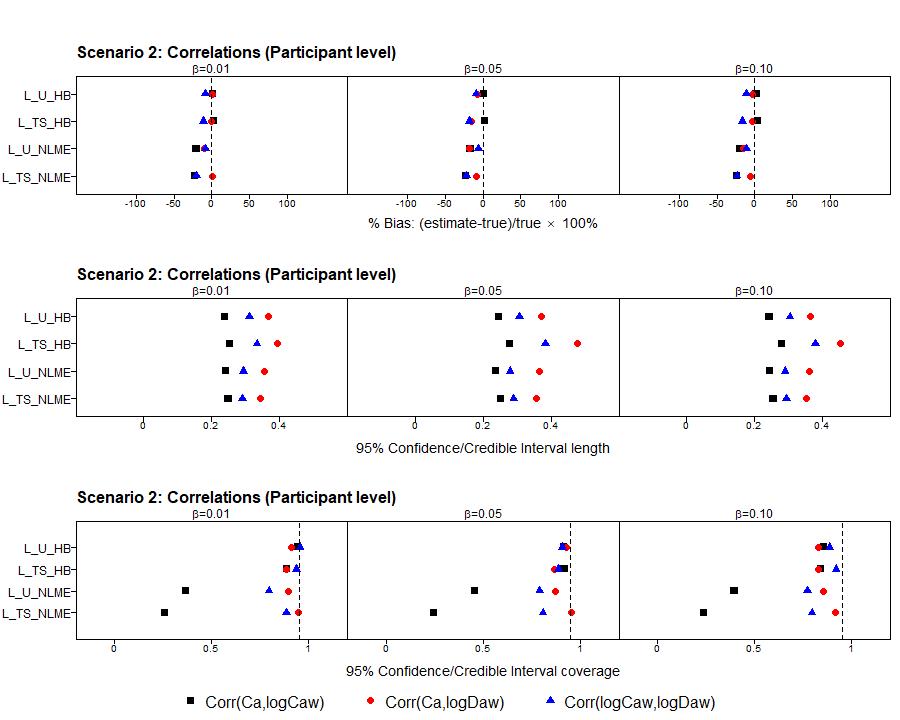

##### B5: Visit level standard deviations σ

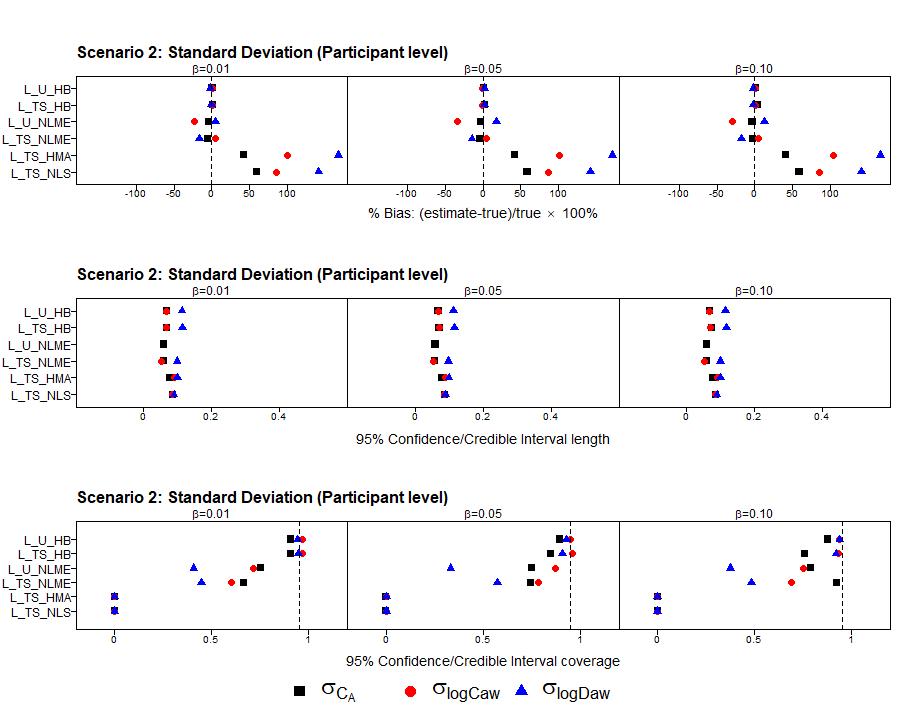

#### C: Scenario 3

| C1: Estimated Associations β (Bias instead of % Bias) |
| --- |
| 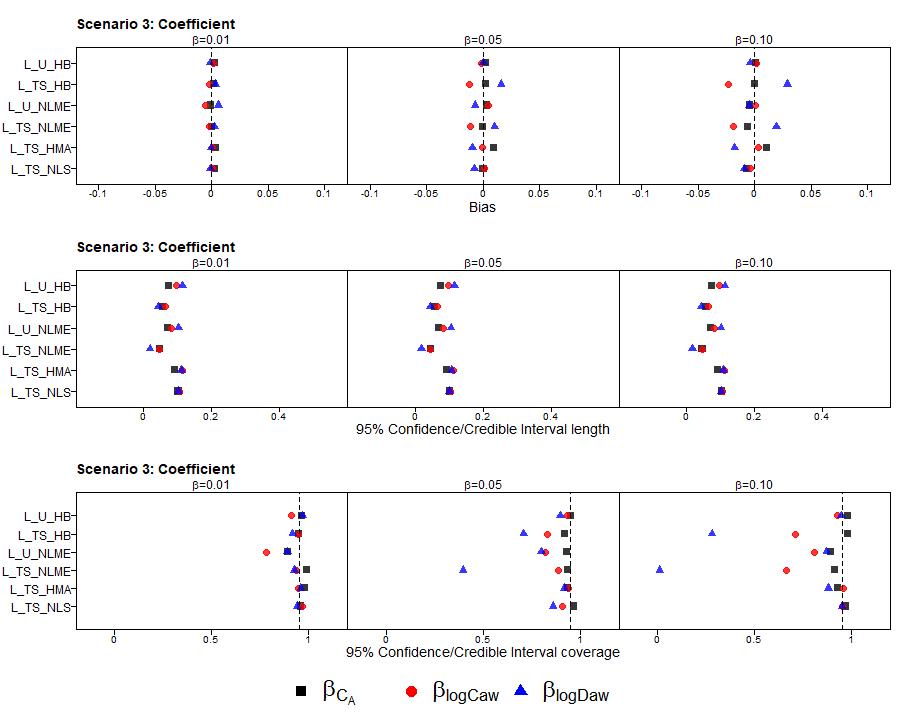 |
| 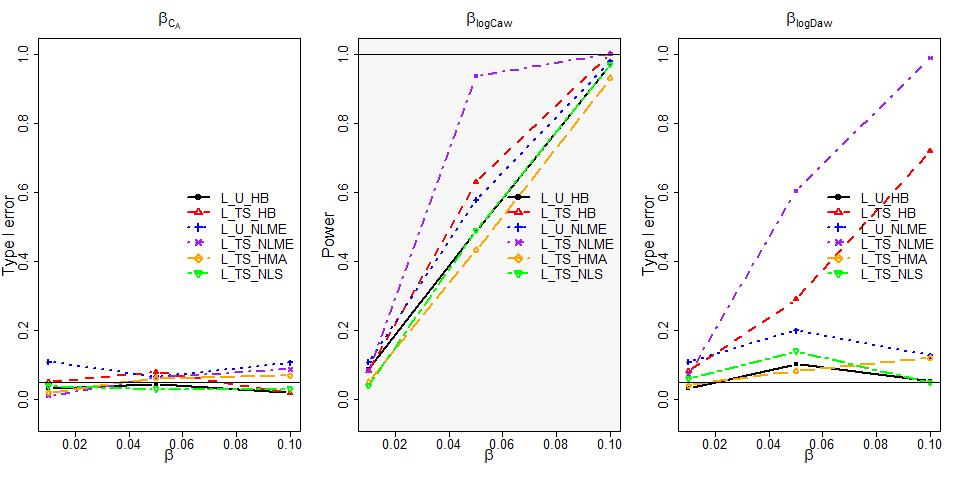 |

##### C2: Population level mean ɑ

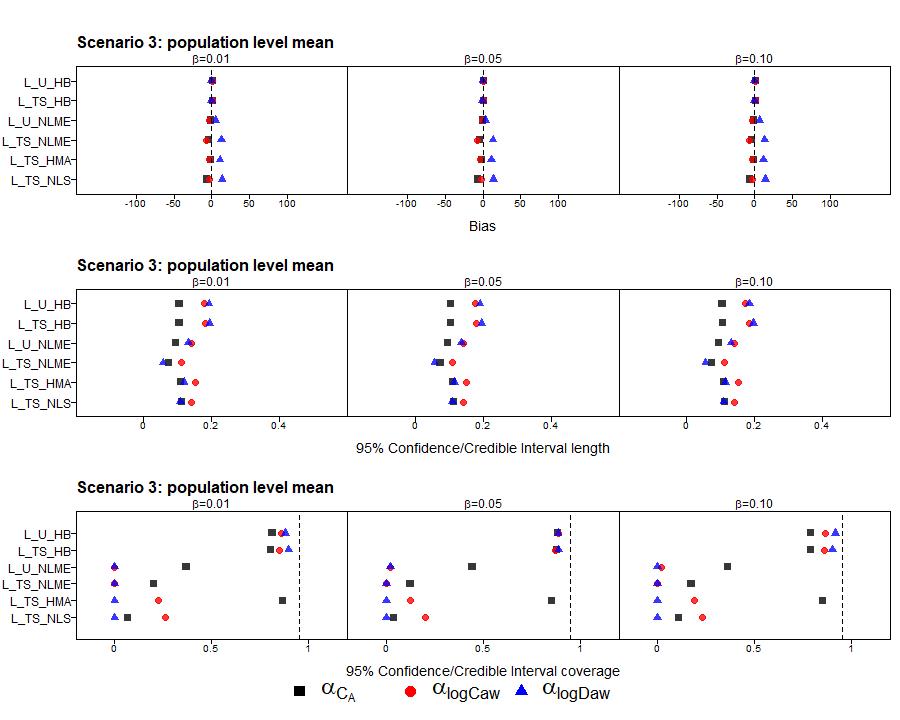

##### C3: Participant level standard deviations τ

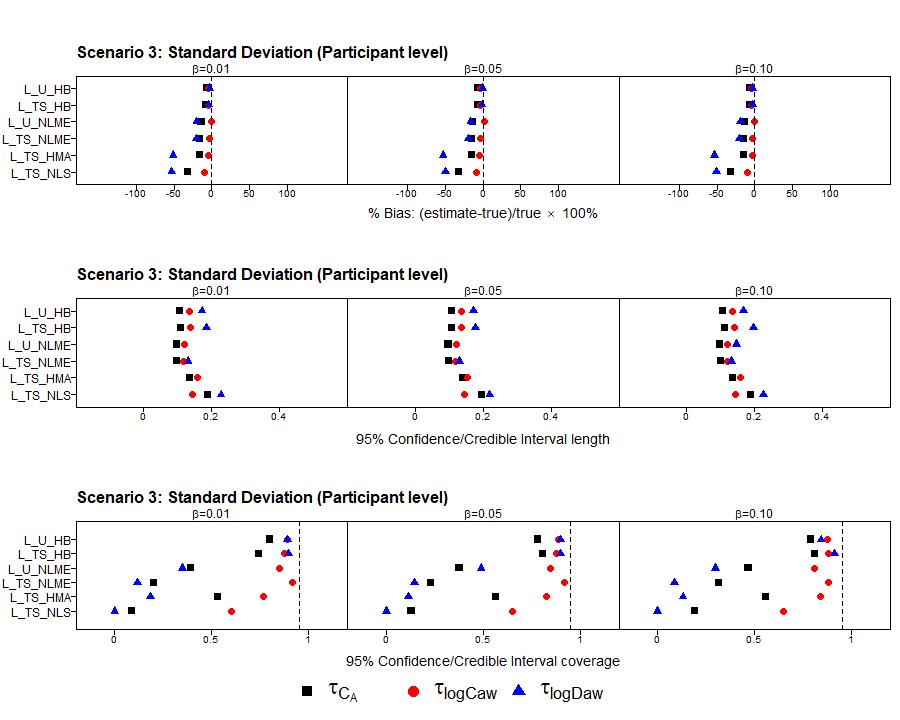

##### C4: Participant level correlations ⍴

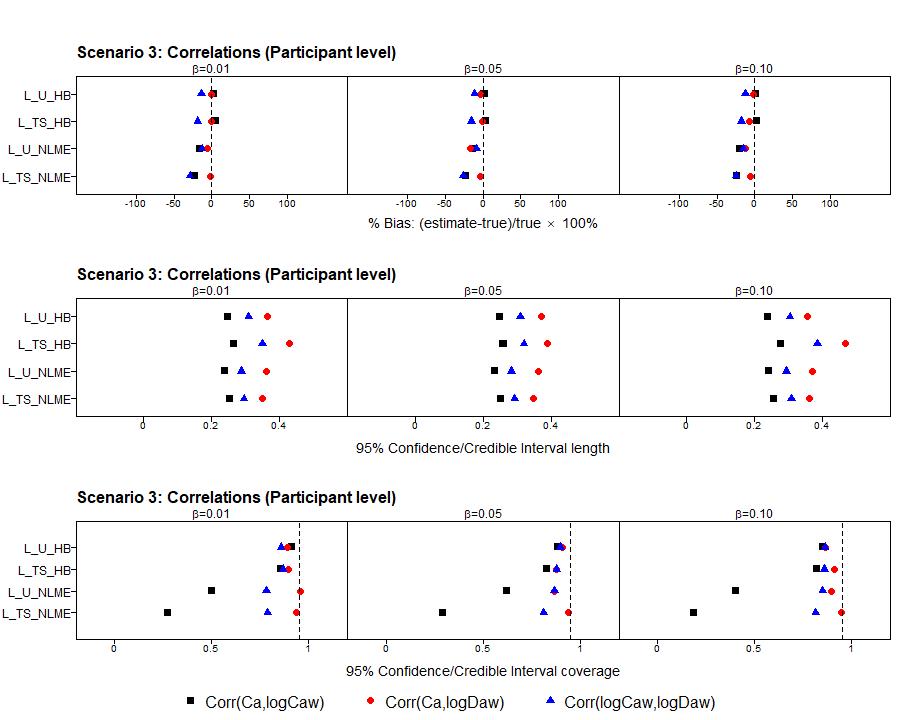

##### C5: Visit level standard deviations σ

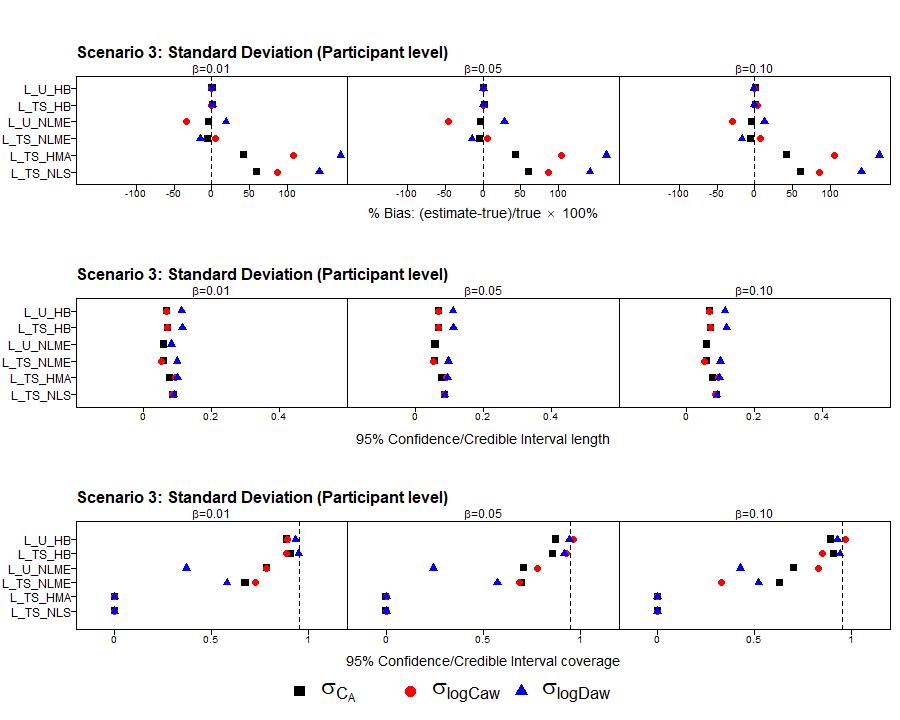

#### D: Scenario 4

| D1: Estimated Associations β (Bias instead of % Bias) |
| --- |
| 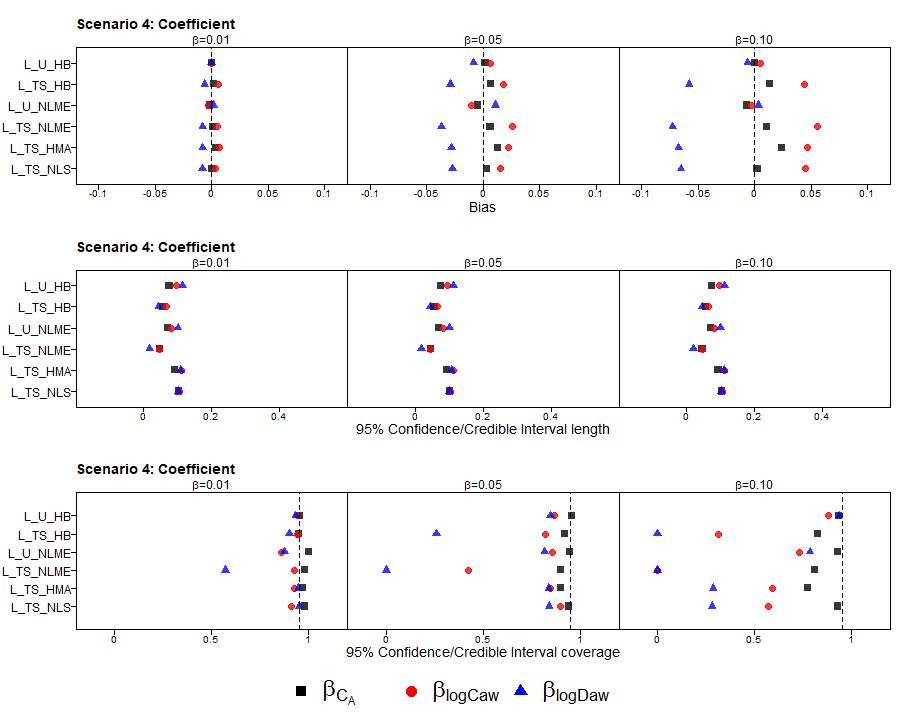 |
| 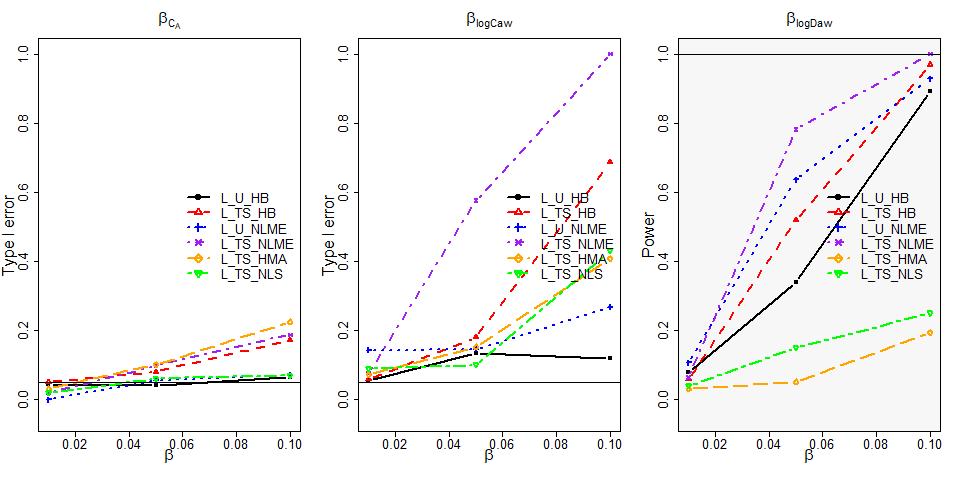 |

##### D2: Population level mean ɑ

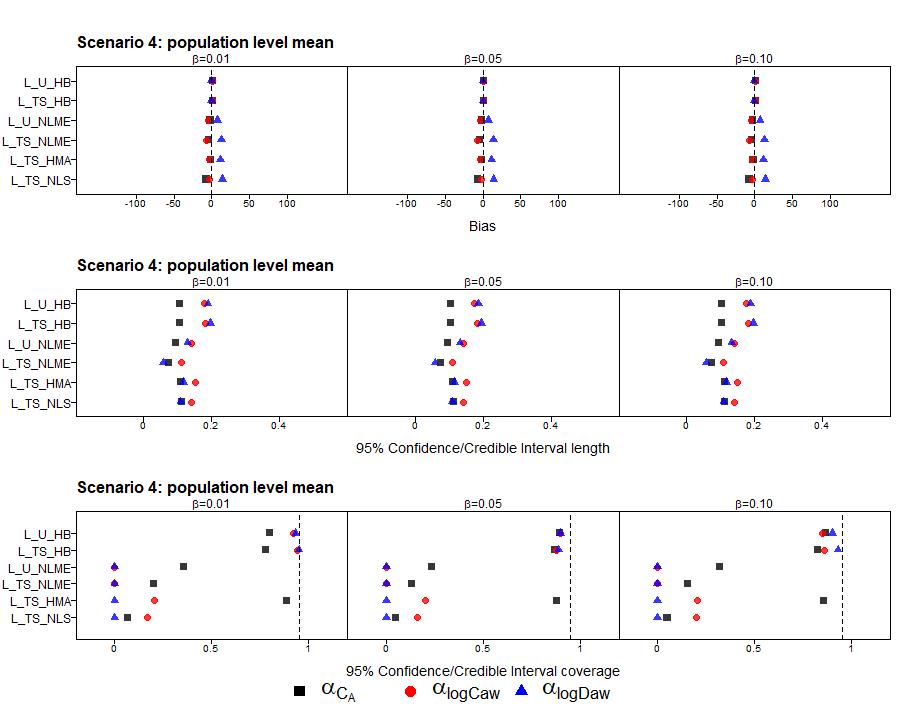

##### D3: Participant level standard deviations τ

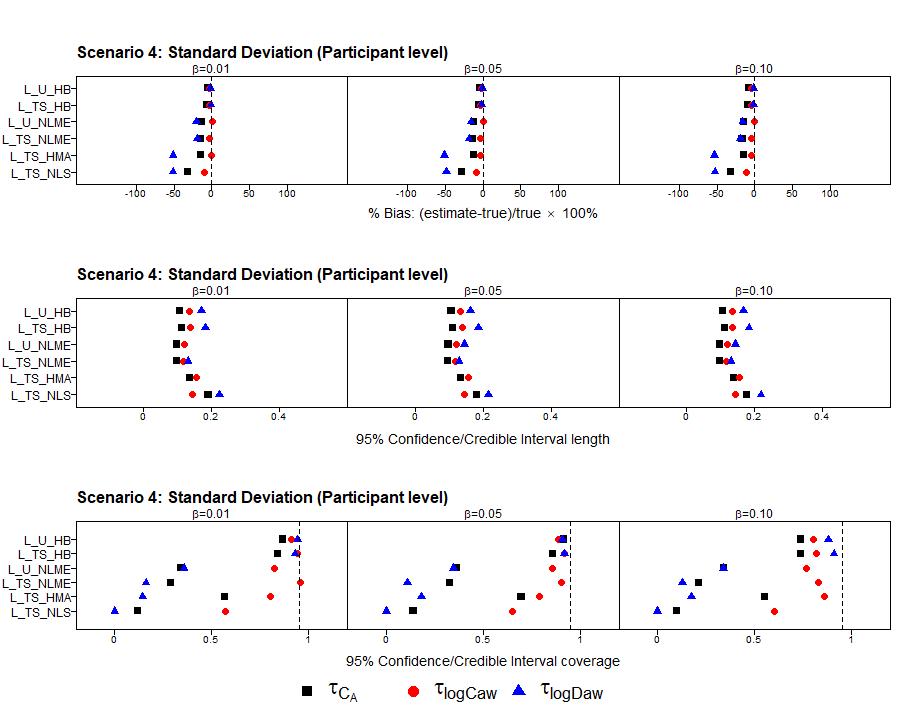

##### D4: Participant level correlations ⍴

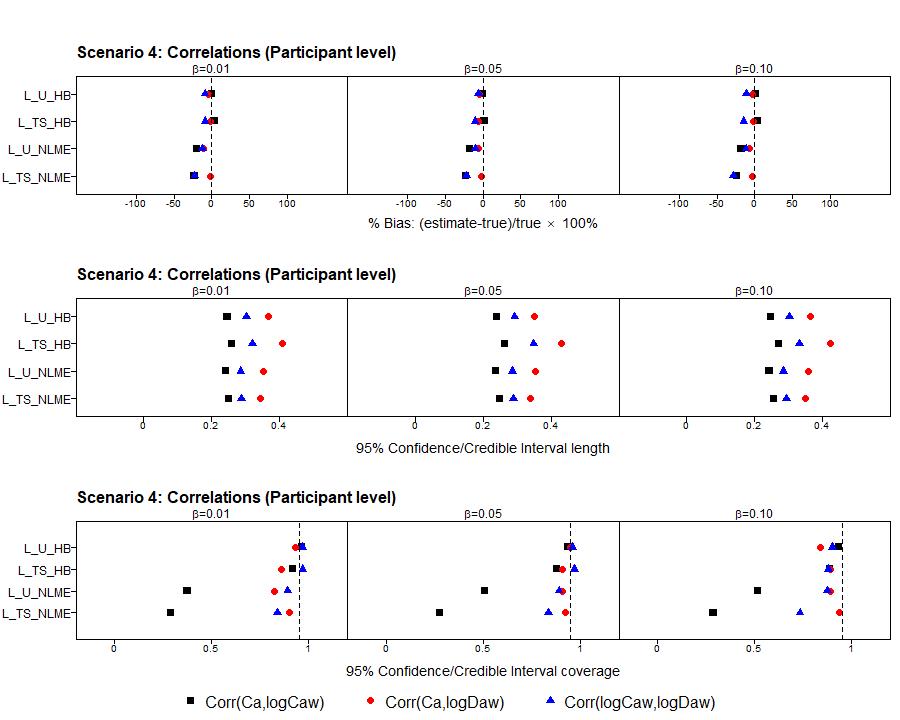

##### D5: Visit level standard deviations σ

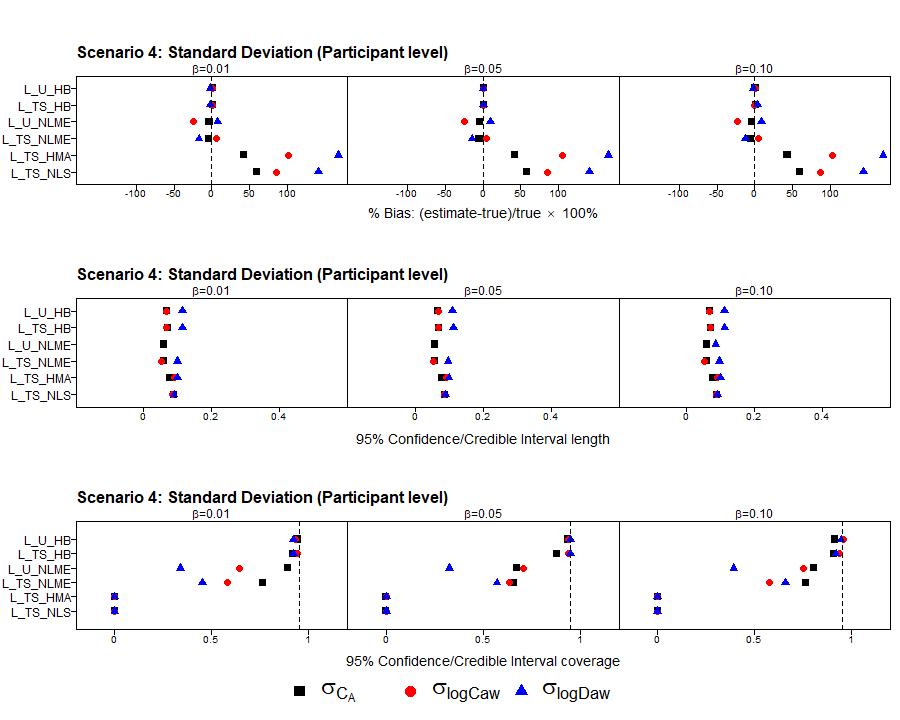

### Figure 2: Summary on all converged datasets (44% - 64%) for Scenario 1 Estimated Associations β^[2]^

Figure 2.a: The percentage bias (% bias), 95 % confidence interval length and the coverage of 95 % confidence intervals

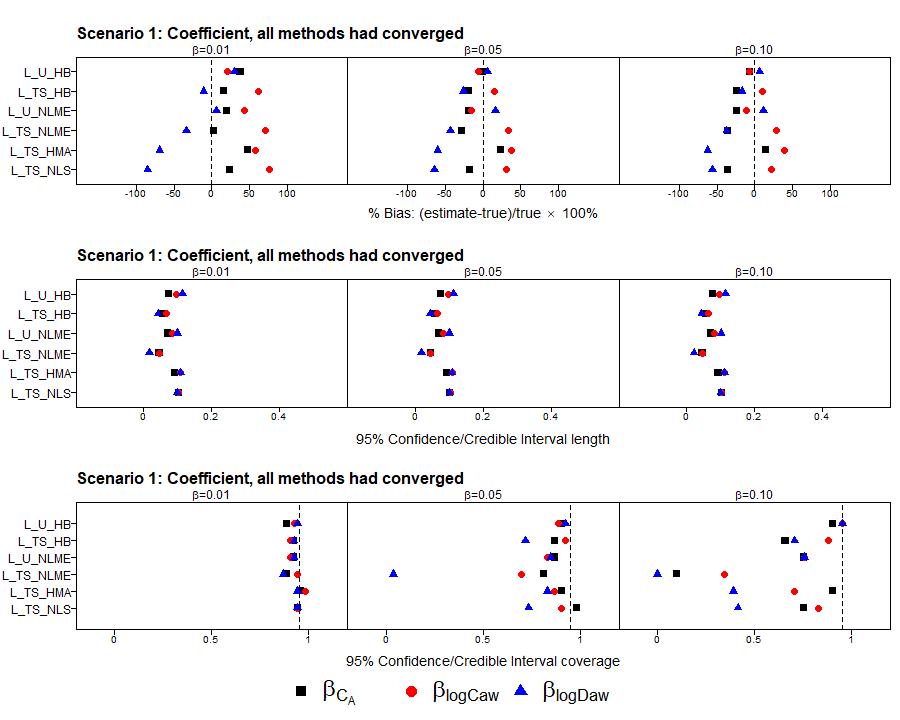

Figure 2.b: The power vs type I error rates:

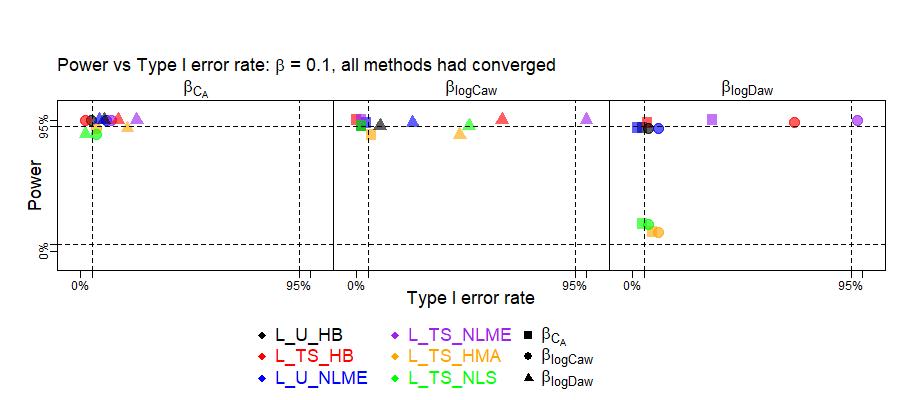

### Figure 3: Constrain the HMA Stage I to be non-negative for NO parameters and ENO.

We previously used all estimations from Stage I for HMA analysis. However, if we use only non-negative estimations, the bias patterns were similar to other two-stage methods but at a cost of 26.4% drop rate on average.

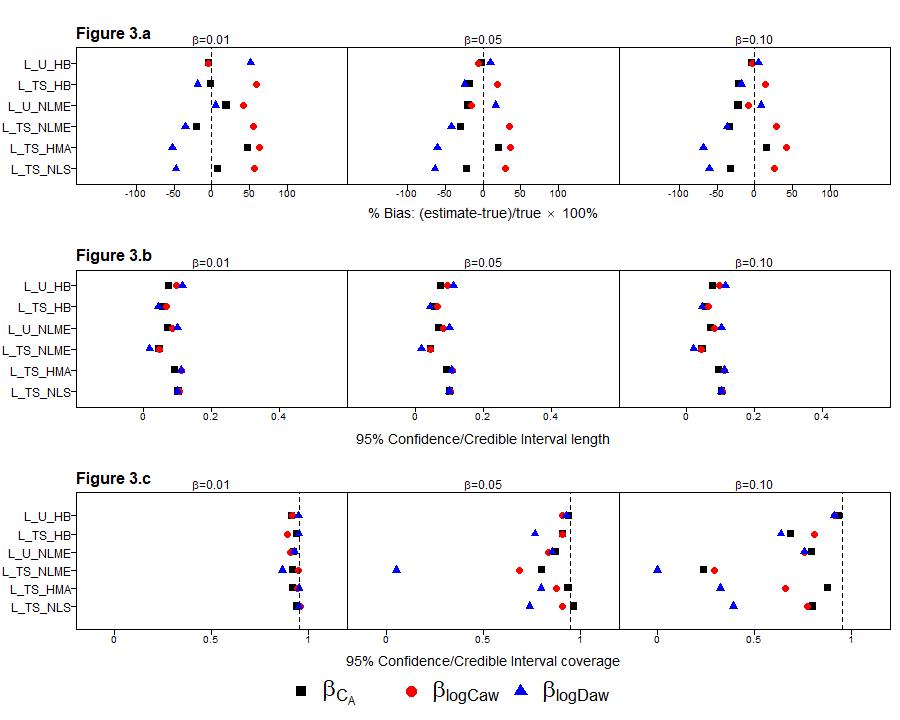

### Table 3: CHS summary

The effects of height to the NO parameters among CHS study data were summarized in Table 3 in the form of Mean (95% CI). Significant associations were highlighted in yellow.

#### Table 3.a: Mean (95% CI) for coefficient of standardized height

| Models | $\beta_{Ca}$ | $\beta_{logCaw}$ | $\beta_{logDaw}$ |
| --- | --- | --- | --- |
| L_U_HB | 0.079 (0.034, 0.125) | 0.158 (0.106, 0.212) | -0.106 (-0.171, -0.044) |
| L_TS_HB | 0.046 (0.014, 0.078) | 0.05 (0.027, 0.072) | 0.006 (-0.015, 0.028) |
| L_U_NLME | 0.092 (0.046, 0.138) | 0.149 (0.104, 0.194) | -0.104 (-0.157, -0.052) |
| L_TS_NLME | 0.038 (0.008, 0.067) | 0.057 (0.033, 0.081) | 0 (-0.021, 0.021) |
| L_TS_HMA | -0.019 (-0.098, 0.06) | 0.063 (0.009, 0.118) | -0.006 (-0.058, 0.046) |
| L_TS_NLS | -0.145 (-1.83, 1.54) | 0.08 (0.033, 0.127) | -0.016 (-0.057, 0.025) |

* On stage I, HMA: 401 failed; NLS: 542 failed

Full results

| parameter | L_U_HB | L_TS_HB | L_U_NLME | L_TS_NLME | L_TS_HMA | L_TS_NLS |
| --- | --- | --- | --- | --- | --- | --- |
| $ɑ_{Ca}$ | 1.943 (1.892, 1.995) | 1.939 (1.902, 1.976) | 1.732 (1.682, 1.782) | 1.726 (1.692, 1.761) | 1.914 (1.831, 1.997) | 1.006 (-0.704, 2.716) |
| $ɑ_{logCaw}$ | 4.412 (4.341, 4.485) | 4.392 (4.359, 4.426) | 4.011 (3.958, 4.064) | 4.004 (3.97, 4.039) | 4.152 (4.095, 4.209) | 4.129 (4.078, 4.18) |
| $ɑ_{logDaw}$ | 2.167 (2.086, 2.246) | 2.187 (2.146, 2.227) | 2.629 (2.569, 2.69) | 2.639 (2.604, 2.674) | 2.574 (2.521, 2.628) | 2.635 (2.592, 2.677) |
| $\tau_{Ca}$ | 0.507 (0.464, 0.557) | 0.528 (0.443, 0.599) | 0.629 (0.588, 0.674) | 0.458 (0.427, 0.491) | NA | NA |
| $\tau_{logCaw}$ | 0.709 (0.653, 0.77) | 0.696 (0.644, 0.754) | 0.52 (0.461, 0.586) | 0.522 (0.497, 0.55) | NA | NA |
| $\tau_{logDaw}$ | 0.861 (0.788, 0.942) | 0.843 (0.776, 0.927) | 0.702 (0.645, 0.764) | 0.546 (0.521, 0.573) | NA | NA |
| $\rho_{Ca,logCaw}$ | 0.816 (0.76, 0.883) | 0.849 (0.773, 0.905) | 0.834 (0.705, 0.91) | NA | NA | NA |
| $\rho_{Ca,logDaw}$ | -0.172 (-0.308, -0.044) | -0.208 (-0.338, -0.058) | -0.448 (-0.561, -0.318) | NA | NA | NA |
| $\rho_{logCaw,logDaw}$ | -0.706 (-0.759, -0.648) | -0.69 (-0.746, -0.631) | -0.571 (-0.633, -0.502) | NA | NA | NA |
| $\sigma_{Ca}$ | 0.659 (0.619, 0.698) | 0.656 (0.615, 0.699) | 0.369 (0.345, 0.394) | 0.468 (0.448, 0.488) | NA | NA |
| $\sigma_{logCaw}$ | 0.359 (0.331, 0.388) | 0.366 (0.339, 0.392) | 0.615 (0.573, 0.66) | 0.304 (0.291, 0.317) | NA | NA |
| $\sigma_{logDaw}$ | 0.37 (0.333, 0.408) | 0.367 (0.331, 0.402) | 0.355 (0.323, 0.39) | 0.243 (0.232, 0.253) | NA | NA |

1. Stage I convergence failure rate is the number of participant’s Stage I models that failed to converge divided by the total number of participants (500)
2. Instead of using all available converged results, we summarize those datasets converged by all methods.
